# Known pathogenic gene variants and new candidates detected in Sudden Unexpected Infant Death using Whole Genome Sequencing

**DOI:** 10.1101/2023.09.11.23295207

**Authors:** Angela M. Bard, Lindsay V. Clark, Erdal Cosgun, Kimberly A. Aldinger, Andrew Timms, Lely A. Quina, Juan M. Lavista Ferres, David Jardine, Elisabeth A. Haas, Tatiana M Becker, Chelsea M Pagan, Avni Santani, Diego Martinez, Soumitra Barua, Zakkary McNutt, Addie Nesbitt, Edwin A Mitchell, Jan-Marino Ramirez

## Abstract

**Purpose:** To gain insights into potential genetic factors contributing to the infant’s vulnerability to Sudden Unexpected Infant Death (SUID).

**Methods:** Whole Genome Sequencing (WGS) was performed on 145 infants that succumbed to SUID, and 576 healthy adults. Variants were filtered by gnomAD allele frequencies and predictions of functional consequences.

**Results:** Variants of interest were identified in 86 genes, 63.4% of our cohort. Seventy-one of these have been previously associated with SIDS/SUID/SUDP. Forty-three can be characterized as cardiac genes and are related to cardiomyopathies, arrhythmias, and other conditions. Variants in 22 genes were associated with neurologic functions. Variants were also found in 13 genes reported to be pathogenic for various systemic disorders. Variants in eight genes are implicated in the response to hypoxia and the regulation of reactive oxygen species (ROS) and have not been previously described in SIDS/SUID/SUDP. Seventy-two infants met the triple risk hypothesis criteria (Figure 1).

**Conclusion:** Our study confirms and further expands the list of genetic variants associated with SUID. The abundance of genes associated with heart disease and the discovery of variants associated with the redox metabolism have important mechanistic implications for the pathophysiology of SUID.

## Introduction

Sudden Infant Death Syndrome (SIDS) is defined as the sudden and unexpected death of an infant younger than one year, for which the cause of death remains unexplained despite a thorough investigation including a complete autopsy, and review of the circumstances of death along with review of the clinical history (American SIDS Institute, 2023). The term SUID or sudden unexpected infant death (SUID) is more encompassing. Using the *International Classification of Diseases, 10th Revision* (ICD-10), both the Centers for Disease Control and Prevention (CDC) and the American Academy of Pediatrics (AAP) define SUID as a larger category that encompasses three types of infant death in children prior to one year of age: “sudden infant death syndrome (SIDS; R95), deaths from other ill-defined or unknown causes (R99), and accidental suffocation and strangulation in bed (W75)” (Lavista Ferres, 2020). SUID remains a significant public health problem. One of the leading causes of infant mortality, it is estimated that SUID contributes to the deaths of approximately 3,400 infants in the United States each year (CDC, 2023). The use of these certified causes of death has varied over time and varies by jurisdiction (Goldstein, 2019).

Many risk factors have been identified for SIDS/SUID including race/ethnicity (CDC, 2023), prematurity (Allen, 2021), maternal smoking during pregnancy (Anderson, 2019; Allen, 2021), lower birth weight (Allen, 2021), bed-sharing (Allen, 2021), prone sleeping (Mitchell, 1991; Mitchell, 2015), recent infection (Goldwater, 2017), geographic variations (Mitchell, 2020), and environmental exposures such as air pollution (Dales, 2004). The recognition of prone sleeping as a major risk factor led to the adoption of safe sleep practices which was associated with significant reductions in SUID rates in the 1990’s and early 2000’s. Increased educational efforts (Mitchell, 1997) could further increase the adherence to safe sleep practices, and also reduce the prevalence of maternal smoking, which in turn could decrease the number of SUID deaths. Unfortunately, despite increased educational efforts, SUID rates have plateaued since the early 2000’s (CDC, 2023).

Further deaths might be prevented through the early recognition of a child that is at higher risk for SUID. DNA sequencing has identified genes related to cardiac, neurologic, and metabolic disorders that could be linked to SUID (Baruteau, 2017; Männikkö, 2018; Tester, 2018a; Tester, 2018b; Gray, 2019; Chahal, 2020; Clemens, 2020; Moore, 2020; Rochtus, 2020; Chahal, 2021; Halvorsen, 2021; Keywan, 2021; Koh, 2022). The early identification of high-risk genes could potentially also be important for preventing mortality and morbidity for children that survive beyond the one-year window for SUID. Identifying risk genes could also help in the diagnosis of a sudden death and could bring closure to surviving members of a family who lost a child. With the aid of genetic counseling, parents could be better informed of the risk of recurrence when considering further children. The knowledge of a genetic vulnerability together with improved recognition devices that monitor multiple physiological parameters, could also increase the usefulness of monitoring devices that have so far failed to prevent deaths (Ramanathan, 2001).

The triple risk model provides an important conceptual framework to assess how a potentially pathogenic gene variant could contribute to a fatal outcome (Guntheroth, 2002). First proposed in 1972, but evolved over time, this concept puts forth the hypothesis that SUID occurs during a critical development period when a vulnerable infant is exposed to an extrinsic stressor (Guntheroth, 2002; Rognum, 1993). A pathogenic gene variant could interfere with any of the three risk factors. It could increase or imbue a child with an intrinsic vulnerability, it could interfere with the response to a particular external stressor, and it could alter the normal developmental trajectory. Importantly, it is generally assumed that none of these isolated threats is significant enough to cause death alone, but when combined, the triple risks reach the threshold for a fatal outcome. Based on numerous neuropathological and neurophysiological studies it is hypothesized that the concurrence of the triple risk (developmental window of post-perinatal age (day 6-364), intrinsic vulnerability and external stressor) leads in most cases to failed arousal to external stressors associated with hypercapnic and/or hypoxic conditions (Garcia, 2013; Kinney, 2019; Nasirova, 2019; Vivekanandarajah, 2021). The conditions become fatal as the child suffers a terminal apnea event leading to severe irreversible hypoxic damage and ultimately cardiac arrest due to the compromised arousal response. However, there are also reports that a child can succumb to apnea, sudden cardiac arrest or perhaps a catastrophic immunological response that is not caused by a failed arousal. Risk genes associated with cardiac and neurologic disorders including cardiomyopathy and channelopathies or metabolic conditions such as fatty acid oxidation or glucose metabolism errors (Baruteau, 2017; Männikkö, 2018; Tester, 2018a; Tester, 2018b; Gray, 2019; Chahal, 2020; Clemens, 2020; Moore, 2020; Rochtus, 2020; Chahal, 2021; Halvorsen, 2021; Keywan, 2021; Koh, 2022) could be potential candidates for such fatal events. While these considerations support the notion that a multitude of pathways can lead to SUID, there are certain features of SUID that are shared and are reproduced world-wide. These common features include male dominance (Allen, 2021), the characteristic age distribution which peaks at the second month and ends at one year of age (Allen, 2021) and maternal smoking (Anderson, 2019) as well as prone sleeping (Mitchell, 1991, 2015).

Next-generation sequencing (NGS) studies have accelerated the study of a wide range of heritable diseases. In the present study we performed WGS of 145 children that succumbed to SUID. Our goals were to (1) validate genes in which altered function have been previously implicated in SUID, and (2) identify novel genes and pathways that tend to be disrupted in SUID so that we can better understand the causes of these deaths. Our complete, non-targeted approach to sequencing enables an unbiased and thorough evaluation of the SUID genome.

## Materials and Methods

## 1. Editorial Policies and Ethical Considerations

Samples obtained from the NIH’s NeuroBioBank (NBB) (https://neurobiobank.nih.gov/) were exempt from the requirement of informed consent because the individuals providing the samples were deceased. However, donor recruitment sites for the NIH NBB typically obtain authorization from the parent/s of the individual. Archived samples obtained from the University of Washington and Seattle Children’s Hospital were granted a waiver for informed consent through Seattle Children’s Research Institute’s Institutional Review Board (IRB), which approved the study. Informed consent was obtained for samples which did not meet criteria for the waiver granted by the IRB. This study is Health Insurance Portability and Accountability Act (HIPAA), General Data Protection Regulation (GDPR) and NIH compliant.

## 2. Affected Infant/Healthy Adult Acquisition

### Affected Infants

#### Source of material

Samples from infants lost to SUID were obtained from three sources: 1. The NIH NBB (n=141, year of death from 1992-2017), 2. Seattle Children’s Research Institute (n=4, year of death from 2010-2014), and 3. University of Washington (n=2, year of death was 2008).

#### Affected infant definition

Autopsy reports and partial medical history were available for only 34 infants, but for 126 infants, some medical examiner notes were included. For the remainder of the cohort, limited data were provided. The cause of death as stated by the ME was accepted. Deaths where there was clear mechanical asphyxia were excluded (n=2). Cause of death of the 145 infants included in the study were SIDS n=89, unexplained/sudden unexplained infant death n=21, undetermined/unknown n=6, sudden unexpected infant death n=11, positional asphyxia n=7, asphyxia/overlay n=11.

#### Postmortem tissue

Tissue was obtained from the 147 infants. For the majority this was brain (95.2%), but for some infants, only liver tissue or blood was available.

### Healthy Adults

Sequencing results (VCFs) from 576 healthy adults were obtained from Veritas (newly acquired by Let’s Get Checked). The control population ranged in age from 18 to 100+, with 47 (8.16%) over 100 years old (Let’s Get Checked, 2023).

See Figure 2 for flow chart.

**Figure 1.**
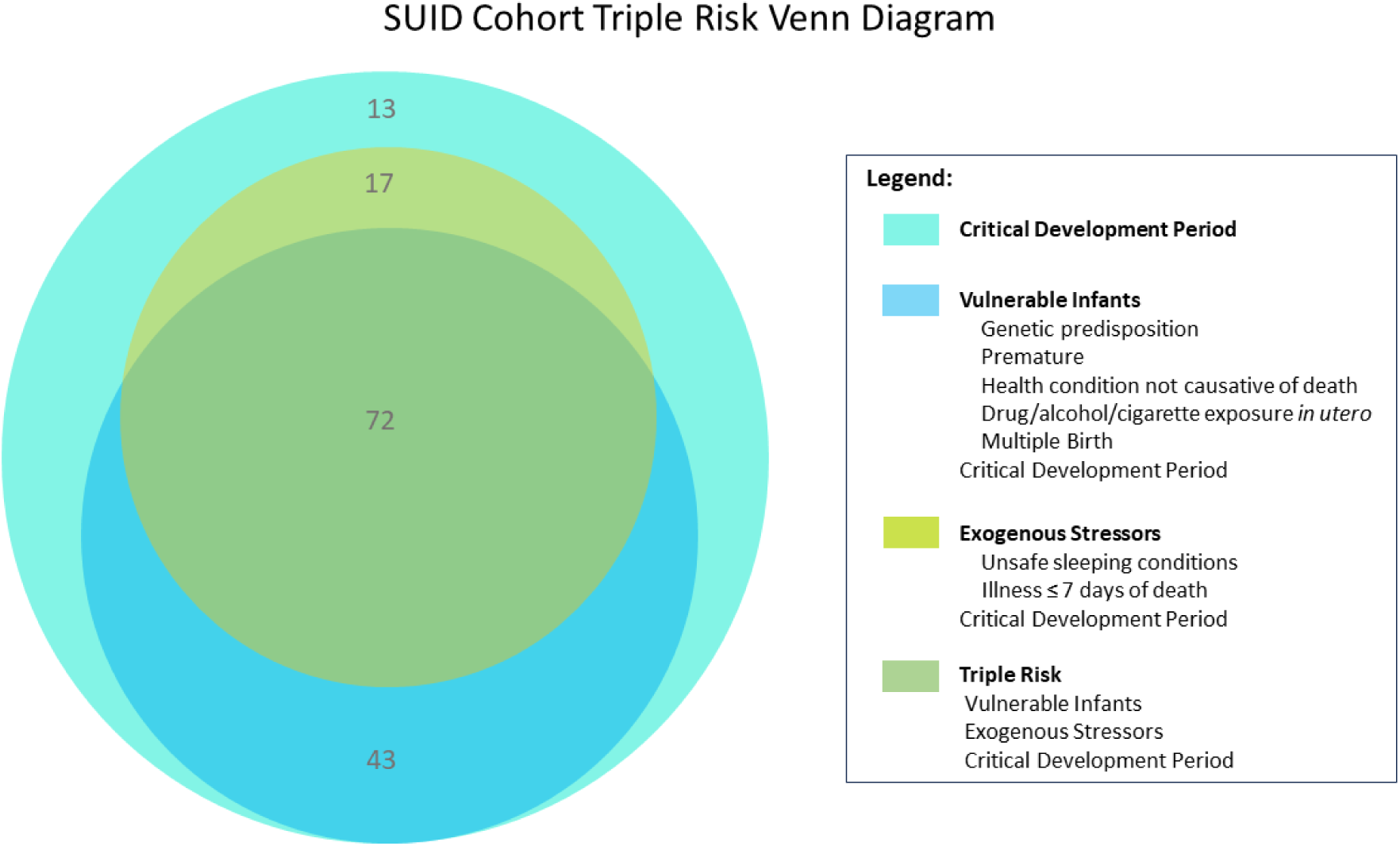
Triple-risk Venn Diagram illustrating the risk factors found among our cohort. Seventy-two infants met the triple risk hypothesis criteria. There were 115 infants determined to be vulnerable. Eighty-nine infants experienced an exogenous stressor at or near the time of death and all 145 infants died during the critical development period, <1 year of age.

**Figure 2.**
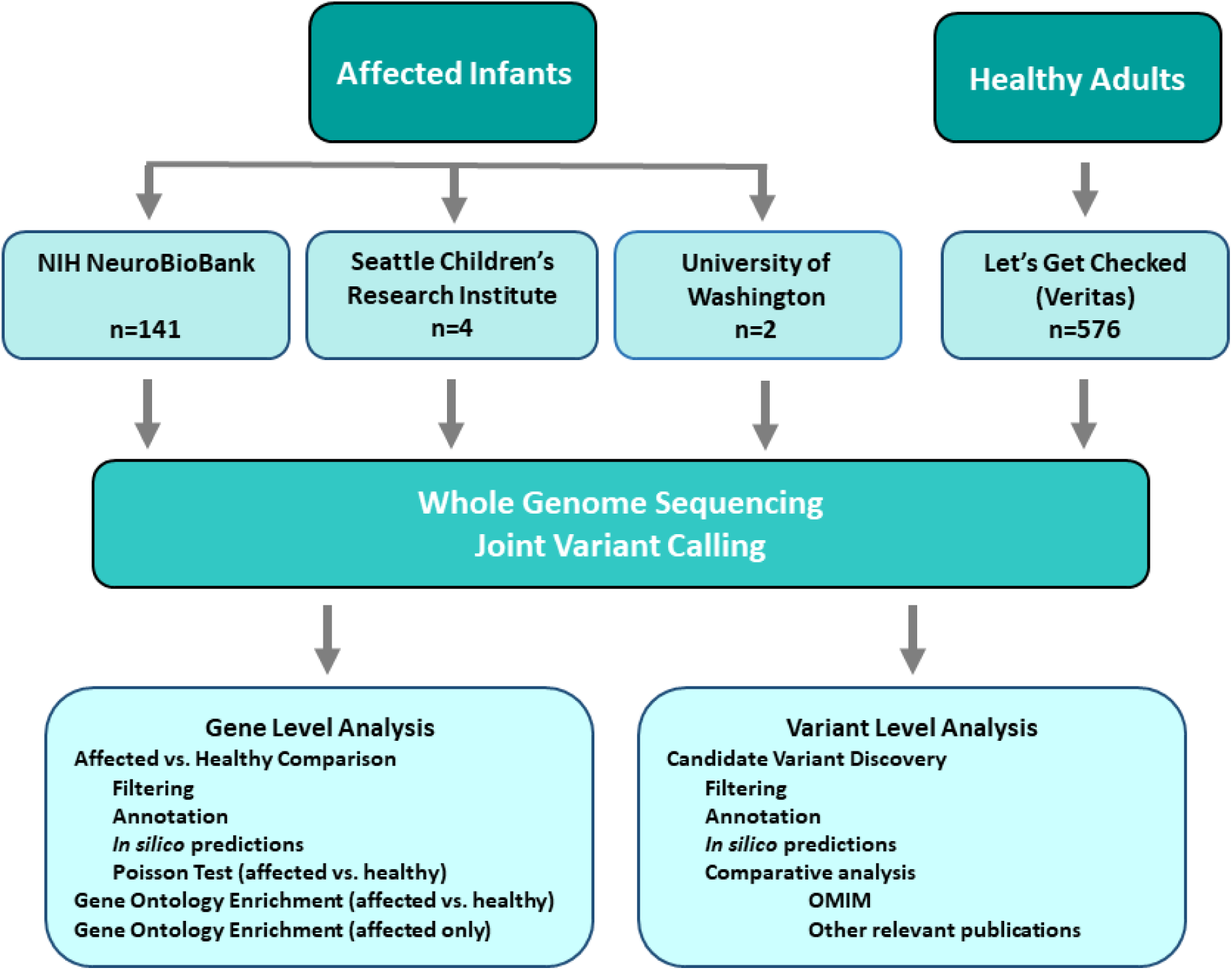
Flow chart. Tissue samples were obtained from 147 SUID affected infants and were submitted for WGS and analysis. VCFs from healthy adults were obtained from Let’s Get Checked (formerly Veritas) and both were joint called for variants. The results were subjected to both gene level and variant level analysis.

## 3. Whole Genome Sequencing

All affected infant and healthy adult samples were subjected to a quality control assessment for high molecular weight DNA and degradation utilizing SNP chip and TapeStation prior to library construction (Let’s Get Checked, 2023). Samples that passed the quality assessment were processed with the TruSeq DNA PCR-free sample preparation kit and sequenced (Let’s Get Checked, 2023). Samples were sequenced to an average of approximately 30X coverage on the Illumina HiSeq X 10 or NovaSeq 6000 next generation sequencers (Let’s Get Checked, 2023). The paired-end sequencing protocol was used targeting an average read-length of 150 base pairs (Let’s Get Checked, 2023). Samples were required to meet or exceed 97% of bases > 10X coverage (Let’s Get Checked, 2023). Veritas performed primary analysis on the results which yielded two FASTQs per sample. Secondary analysis was also performed by Veritas using Microsoft accelerated algorithms and yielded one BAM file and genomic variant call file (gVCF) per sample. Tertiary analysis was completed at Microsoft and SCRI. Joint genotype calling was performed with GATK GenotypeGVCFs.

## 4. Population Structure Analysis

To confirm a lack of major batch effects between affected infants and healthy adults, population structure was visualized across the 721 individuals. The joint-called variant call file (VCF) was pruned with BCFtools (Li, 2009) to only retain variants with less than 10% missing data, removing variants with linkage disequilibrium of 0.25 or greater with another variant, retaining a maximum of 10 variants per 100 kb. The variants were then imported into R and further filtered to only retain autosomal, biallelic variants with excess heterozygosity below 125 and an alternative allele frequency between 0.02 and 0.5, yielding 9252 variants. Principal components analysis, using the PPCA method from the pcaMethods Bioconductor package to handle missing data, was run on the genotype matrix (Stacklies, 2007). The first 50 axes were retained and used as input to UMAP (McInnes, 2018) for visualization. Self-identified race and results from GrafPop (Yumi, 2019) were used to determine the ancestry of each cluster (Figure 3).

**Figure 3.**
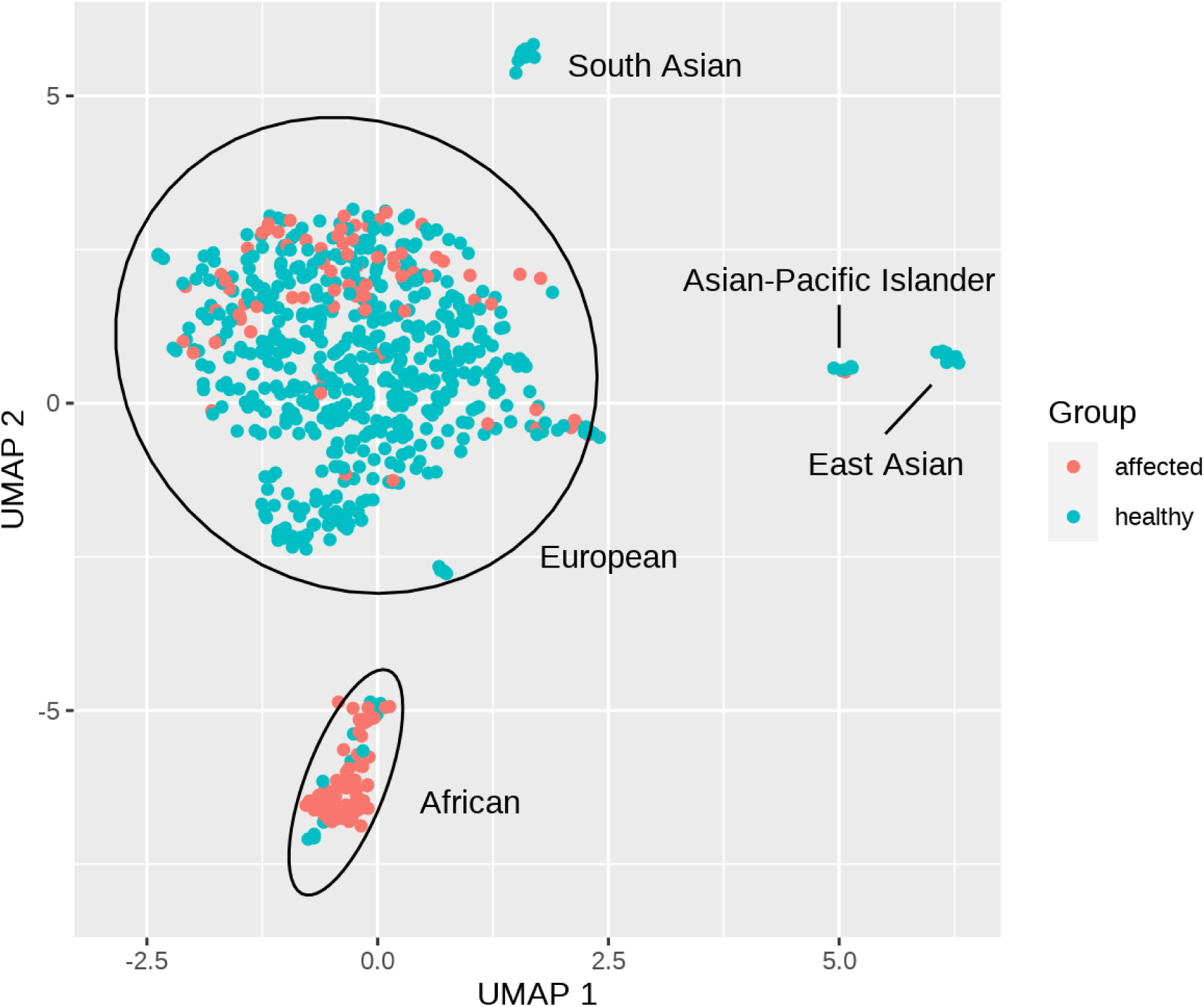
UMAP to visualize population structure across affected infants and healthy adults. Ancestry of clusters is labeled based on self-identified race as well as GrafPop results on the genotype data.

## 5. Gene Level Analysis

### 5.1. Annotation and filtering

BCFtools (Li, 2009), SnpEff (Cingolani, 2012a), Slivar (Pedersen, 2021), ANNOVAR (Wang, 2010), and SnpSift (Cingolani, 2012b) were used to add functional annotation, predictions of pathogenicity, gnomAD allele frequencies (Karczewski, 2020), and data from ClinVar (Landrum, 2013) to the VCF. Variants were excluded if they were listed as Benign or Likely benign in ClinVar, if their inbreeding coefficient was below -0.4 or above 0.4, if their excess heterozygosity was above 125, or if the alternative allele frequency was greater than 0.05 within the dataset. To eliminate variants with dramatically different call rates between affected and healthy, Fisher’s exact test was performed for each variant on the contingency matrix of missing vs. called genotypes in affected and healthy, and variants were excluded if the odds ratio was greater than 8 or less than 0.125 and the P-value was less than 0.01. An allele frequency cutoff for maximum frequency in gnomAD was set to 0.0001 for putative dominant variants, and 0.005 for putative recessive variants. Two allele frequency filters were then applied. (1) In the first filter, if a variant was homozygous in any individual, it had to pass the recessive allele frequency threshold in all populations. Other variants had to pass the dominant allele frequency threshold in all populations. This filter yielded 18,498 variants. (2) The second filter was the same as the first, except that variants had to be completely absent in the healthy adults (unless they were homozygotes in the affected and only heterozygotes in the healthy), yielding 2764 variants.

Three filters, decreasing in stringency, were then applied to these two filtered datasets. (1) The first filter retained only variants labeled as Pathogenic (P) or Likely pathogenic (LP) in ClinVar, yielding 120 variants across 102 genes out of the 18,498 variants in affected infants and healthy adults, and 21 variants across 21 genes out of the 2764 variants only in affected infants. (2) The second filter included all variants from the first, plus any that were indicated as deleterious using three out of six computational methods, including MetaSVM, FATHMM or fathmm_MKL_coding, Polyphen2 HDIV or HVAR, CADD, GERP++, and DANN. This filter yielded 3474 variants across 2673 genes out of the 18,498 variants in affected and healthy, and 656 variants across 633 genes out of the 2764 variants only in affected infants. (3) The third filter included all variants from the first two plus any splicing, frameshift, or stop gain variants. This filter yielded 9764 variants across 5030 genes out of the 18,498 variants in affected and healthy, and 1151 variants across 988 genes out of the 2764 variants only in affected infants.

### 5.2. Gene-level comparison of affected infants and healthy adults

The three filtered sets of variants found across affected and healthy (120 for ClinVar P/LP, 3474 for ClinVar P/LP or computational methods, and 9764 for ClinVar P/LP, computational methods, frameshifts, splicing, or stop gain) were used to test for genes in which variants were more frequent in affected infants than in healthy adults. For each variant set, a gene-by-individual matrix was constructed, tallying the number of variants within each gene and individual. Variants that were homozygous in any individual were only counted in individuals in which they were homozygous. A baseline rate ratio was also calculated for each variant set as

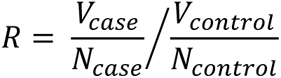

where *V* is the sum of all observations of variants across all genes within the set, and *N* is the total number of genotype calls. For each gene with at least five observations of variants in a given variant set, a one-tailed Poisson test was then run to see if the rate ratio within that gene was higher than the baseline rate ratio. Multiple testing correction was performed across genes within each variant set using the False Discovery Rate (FDR) method of Benjamini and Hochberg (Benjamini, 2001). See Figures 4 and 5.

**Figure 4.**
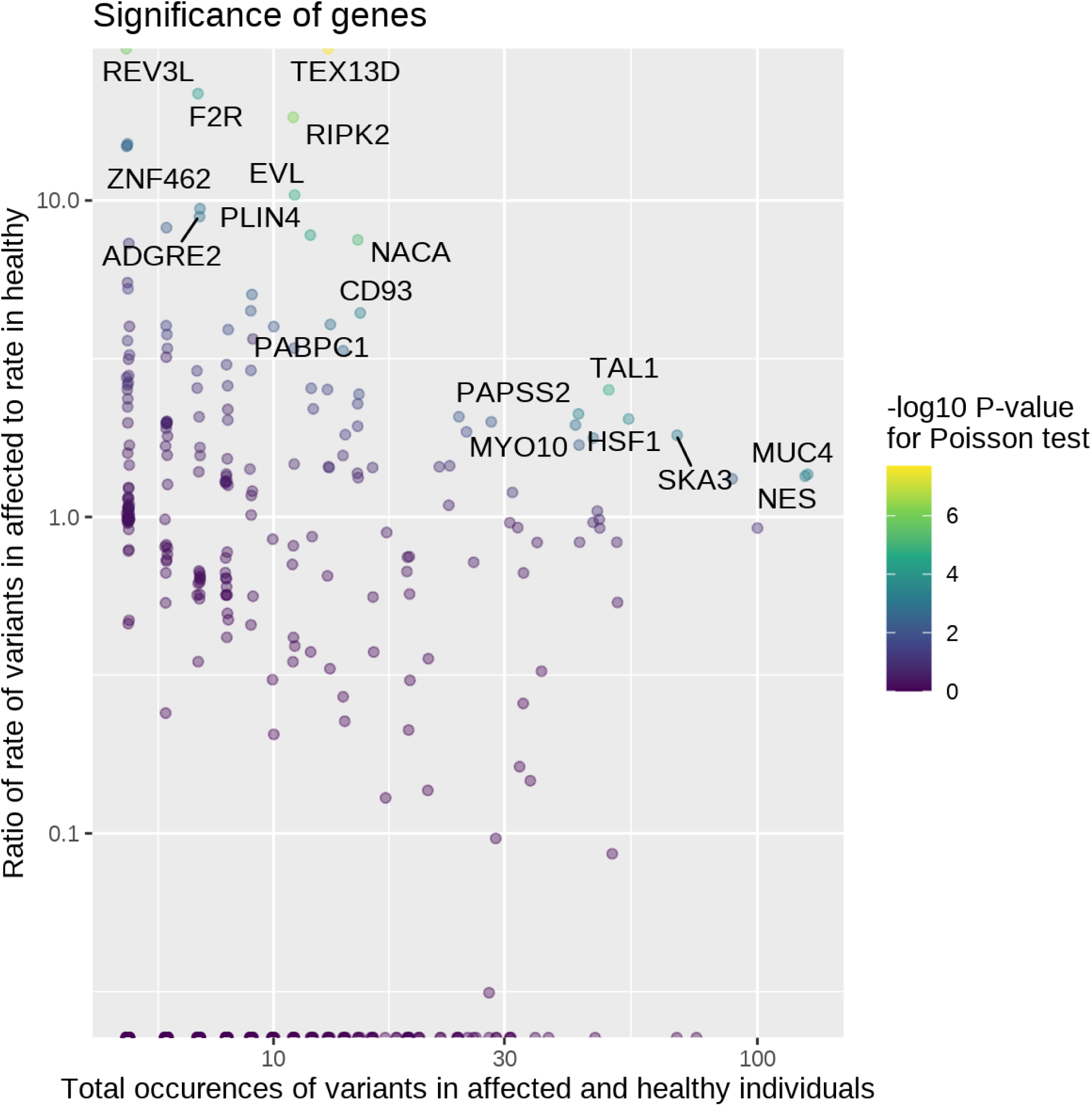
Gene-level Poisson test for rate of observation of variants in affected infants vs. healthy adults. A total of 9764 variants filtered for allele frequency and evidence of pathogenicity were included for testing. Genes with significance at FDR < 0.05 are labeled.

**Figure 5.**
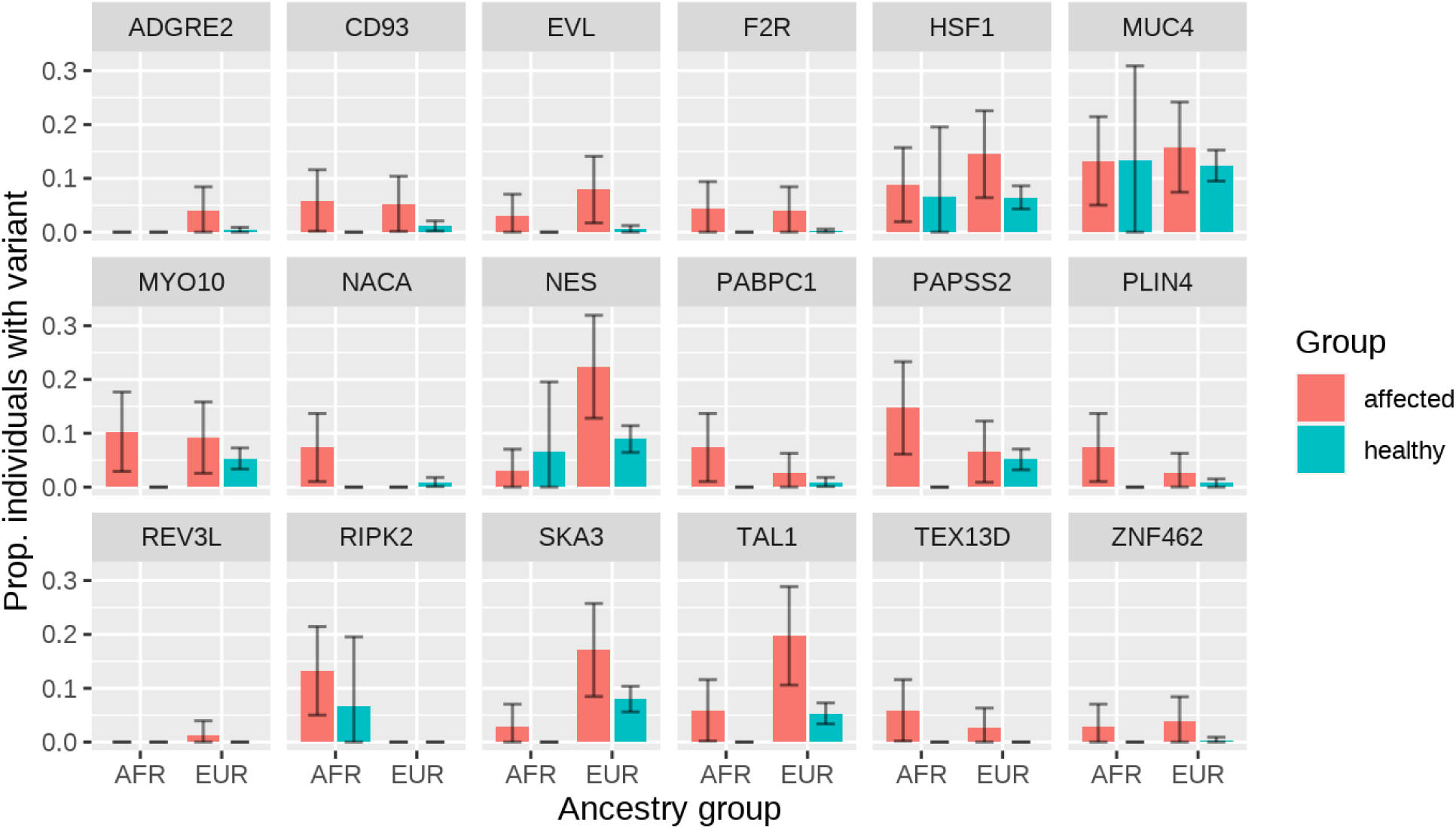
Proportion of individuals, in each of 18 genes with a significantly higher rate of variants in affected infants as compared to healthy adults, with at least one potentially pathogenic variant. Ancestry was determined by clustering in UMAP. Individuals are split by affected/healthy and by ancestry group, with individuals of Asian ancestry not shown since only one affected infant was of Asian ancestry. AFR = African ancestry; EUR = European ancestry. Error bars indicate 95% margin of error on proportions. A total of 9764 variants filtered for allele frequency and evidence of pathogenicity were analyzed.

### 5.3. Gene ontology enrichment in affected infants vs. healthy adults

A Poisson test similar to that used to detect significant genes was run on 13,774 gene ontology (GO) terms, using the sum of observation of variants across genes within each ontology term. Multiple testing correction was not appropriate given overlap among ontology terms, so instead a p-value cutoff of 0.00001 was used.

### 5.4. Gene ontology enrichment in affected infants only

Lists of genes in affected infants only for each of the three sets were analyzed in STRING-DB for protein – protein interactions and enrichment in GO terms (Szklarczyk, 2019). Genes from the three sets were analyzed using the full STRING network. The required score was set to the medium confidence level of 0.400 and FDR stringency was set to medium, 5%.

## 6. Variant Level Analysis: Gene Discovery Mendelian Pipeline

The control VCF was decomposed, subset and annotated with an Ensembl gene annotation using bcftools (Let’s Get Checked, 2023). Slivar was used to prioritize variants based on quality, coverage, allele balance and population frequency in gnomAD (Karczewski, 2021; Pedersen, 2021). In addition, Slivar was used to add healthy adult counts and gene-based information for probability of being loss-of-function intolerant (pLI) score and any Online Mendelian Inheritance in Man (OMIM) associations (Pedersen, 2021; OMIM, 2022). Annovar was used to add additional variant-based annotation including: refGene model, ClinVar, CADD, GERP++, PolyPhen2 score and Geno2MP information (Wang, 2010).

Variants were prioritized using the following criteria: *≥*10 reads; genotype quality *≥*15; allele balance of >0.8 for homozygous alternate and between 0.2 and 0.8 for heterozygous; max gnomAD population frequency of *≤*1%, and were determined to be functional (including missense, frameshift, or splicing [splice acceptor, splice donor or splice region i.e. within 8bp of a exon]). Variants that appeared in healthy adults were excluded.

Included here are variants only described as rare/ultra-rare (<0.005%) in gnomAD, with noted exceptions, associated with autosomal dominant (AD) conditions, though some variants are also associated with multiple conditions including those of autosomal recessive (AR) inheritance. Infants were all heterozygous for the variant unless otherwise stated. When a variant is associated with AR conditions alone, it is only included if the infant had two variant alleles for that gene in different phases. The allele frequency of these variants is reported specific to the race/ethnicity of the infant. When the race/ethnicity of the infant was not available, the total allele frequency for the combined racial/ethnic groups in gnomAD is reported (Karczewski, 2021).

## RESULTS

## 1. Characteristics Cohort

The characteristics were similar to that reported previously (3). 51.2% of infants were white and 44.7% were black. 54.5% were boys. The median age at death was 86 days (IQR = 59, 110). There were 10 twins and 1 triplet (Table1).

As the available data were incomplete it is difficult to report the true frequency of other factors. However, there were 30 infants, 20.7% of the cohort, reported to have had mild illness at or within one week of death. Mostly these illnesses related to the respiratory tract, however, these were not determined to be the cause of the death. 41 (38.7% of those reporting co-sleeping) were co-sleeping with parents and/or siblings, however for 39 (26.9% of the cohort) it was not known whether they were co-sleeping.

Infants were determined to be vulnerable if they had at least one of the following risk factors: prematurity, genetic predisposition (based on WGS results), exposure to cigarettes, alcohol and/or drugs *in utero*, having a health condition not determined to be causative of death, or being a twin or triplet. One hundred fifteen (79.3% of the cohort) met these criteria. Eighty-nine infants (76.7%) were described as experiencing an exogenous stressor at or near their time of death. The following risk factors were considered to be exogenous stressors in this study: illness within seven days of death, co-sleeping, prone sleeping, or other unsafe sleep such as excess bedding. All infants in the study died during the critical development period, between one week and one year of age. Risk factors among our cohort as they occur within the triple risk hypothesis categories, vulnerable infant, exogenous stressor, and critical development period are illustrated in Figure 1. Many infants had more than one risk factor in multiple categories, with no clear association between vulnerabilities, exogenous stressors, and age at death (Figure 6).

**Figure 6.**
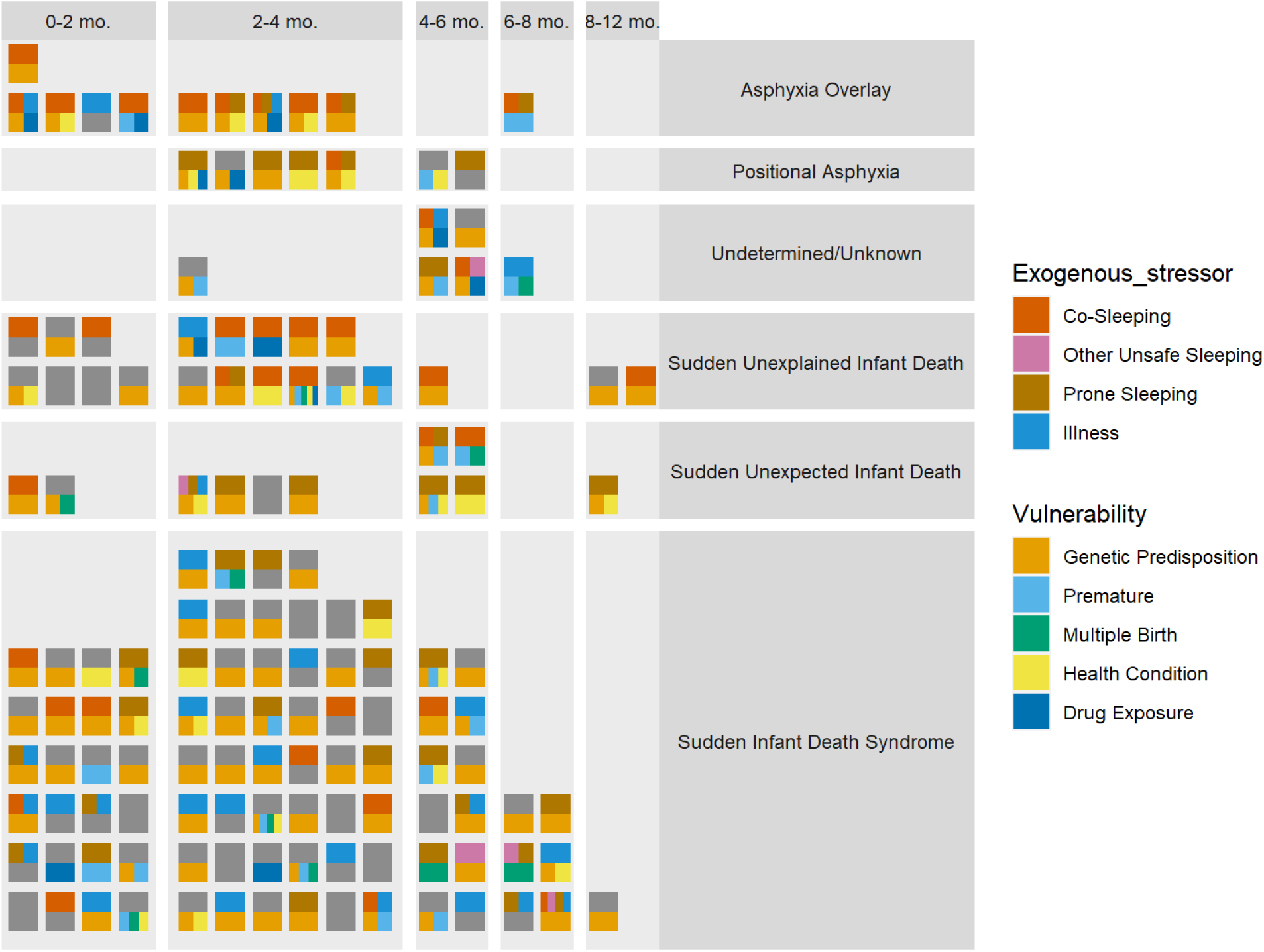
Visual representation of risk factors, cause of death and age at death for 145 infants in the study cohort. Colors on the top of each block indicate exogenous stressors, while colors on the bottom indicate vulnerabilities. Grey indicates no exogenous stressors and/or vulnerabilities. Cause of death is listed as stated by the medical examiner. “Genetic predisposition” indicates that the infant had at least one of the variants highlighted in our study.

## 2. Population Structure Analysis

All major clusters contained both affected and healthy individuals, indicating a lack of confounding batch effects, although ancestry proportions differed between affected and healthy as expected, with affected infants having a much higher proportion of African-Americans (Figure 3). Based on clustering in UMAP, self-reporting of race, and ancestry inference using GrafPop, individuals were divided into three broad ancestry groups: Asian (one affected infant and 35 healthy adults), African (68 affected and 15 healthy), and European (76 affected and 526 healthy).

## 3. Gene Level Analysis

### 3.1. Genes significantly enriched for functional variants in affected infants vs. healthy adults

Using 9734 variants that passed the least stringent filtering criteria (P/LP on ClinVar, predicted pathogenic with at least three computational methods, and/or a frameshift, splice site, or stop gain variant), 18 genes were significant at FDR < 0.05 for having a higher affected:healthy ratio for rate of variants than the baseline ratio of 0.645 (Figure 4, Supplemental Table S1). For some of these genes, affected infants were more likely to have a variant than healthy adults regardless of ancestry, whereas others only showed potentially pathogenic variants in affected infants of European or African ancestry (Figure 5). Significant genes were largely driven by frameshift and splicing variants that were more common in affected than healthy (Supplemental Table S2).

### 3.2. Gene ontology enrichment in affected infants vs. healthy adults

Using the same 9734 variants from the gene-level analysis, 33 gene ontology terms had *P* < 0.00001 for the affected:healthy ratio of rate of variants exceeding the baseline ratio of 0.645. Out of these 33 terms, 17 had lower P-values than any of their constituent genes (Supplemental Table S3). Notable terms among those 17 include Golgi lumen (GO:0005796) and maintenance of gastrointestinal epithelium (GO:0030277) driven by variants found across multiple mucin genes; positive regulation of cysteine-type endopeptidase activity involved in apoptotic process (GO:0043280) driven by *F2R* and *HSF1*; positive regulation of mitotic cell cycle (GO:0045931) driven by *TAL1* and *HSF1*; and several involving *RIPK2* including JUN kinase kinase kinase activity (GO:0004706), response to exogenous dsRNA (GO:0043330), positive regulation of T-helper 1 cell differentiation (GO:0045627), response to interleukin-1 (GO:0070555), response to interleukin-12 (GO:0070671), and cellular response to muramyl dipeptide (GO:0071225).

### 3.3. Gene ontology enrichment in affected infants only

Submission of the first set of 21 genes to STRING which were found with variants in affected infants only, and previously reported as P or LP in ClinVar revealed no significant enrichment in any GO terms (Benjamini, 2001). Submission of the second set of 633 genes that included the previous 21 genes in addition to genes containing variants predicted to be deleterious by at least 50% of the computational methods applied which revealed significant enrichment in 140 GO terms at FDR < 0.05 (including 17 GO terms with FDR ≤ 0.0001), with a protein-protein interaction P-value < 1.0e-16 indicating more connectivity in the network than would be expected from a random set of genes (Supplemental Table S4). Of the 17 most significantly enriched terms, notable enrichments included actin filament binding (GO:0051015), ATP-dependent activity (GO:0016887), extracellular matrix structural constituent (GO:0005201), cytoskeletal motor activity (GO:0003774), and microfilament motor activity (GO:0000146), all by a factor of >2.5. Additionally, 8 GO terms were enriched by a factor of >5 with a significance of P ≤ 0.01. Among them, axonemal dynein complex (GO:0005858), intracellular ligand-gated ion channel activity (GO:0005217), myosin complex (GO:0016459), stereocilium (GO:0032420), cardiac cell development (GO:0055006), cardiac muscle cell differentiation (GO:0055007), dynein complex (GO:0030286), and DNA helicase activity (GO:0003678). Another 4 GO terms were enriched by a factor of 10 including ryanodine-sensitive calcium-release channel activity (GO:0005219), collagen-activated tyrosine kinase receptor signaling pathway (GO:0038063), collagen-activated signaling pathway (GO:0038065), and minus-end-directed microtubule motor activity (GO:0008569), though not as significantly (P ≤ 0.05).

Submission of the third set of 988 genes to STRING which were found with variants in affected infants only, previously reported as P/LP in ClinVar, predicted by our computational methods to be deleterious and including any frameshift, stop-gained or splicing variants revealed significant enrichment in 91 GO terms at FDR < 0.05 (with 16 at FDR ≤0.0001), with protein-protein interaction significant at P < 1.1e-14 (Supplemental Table S5) (Benjamini, 2001). Of the 16 most significant GO terms, notable enrichment included actin filament binding (GO:0051015), by a factor of >3. An additional seven GO terms were enriched by a factor of 6 including microfilament motor activity (GO:0000146), actin-dependent ATPase activity (GO:0030898), spectrin binding (GO:0030507), calcium-release channel activity (GO:0015278), intracellular ligand-gated ion channel activity (GO:0005217), ligand-gated calcium channel activity (GO:0099604), and myosin complex (GO:0016459), though not as significantly (P<0.05).

## 4. Variant Level Analysis

### 4.1. Variants found in affected infants only

We performed a comparative analysis between variants in our cohort and variants in genes found in other relevant studies (Tester, 2018b; Gray, 2019; Halvorsen, 2021; Koh, 2022) as well as Online Mendelian Inheritance in Man (OMIM) terms associated with SIDS and Death in Infancy (OMIM, 2022).

We report here 156 variants of interest in 92 infants (63.4% of the cohort) in 86 genes, including 128 rare/ultra-rare variants in 71 genes that have been previously associated with SIDS/SUID/SUDP. The majority of these variants, 115, were missense variants. Twelve variants occurred in the splice region as well as five splice acceptor and two splice donor variants. Ten variants resulted in a premature stop and nine resulted in a frameshift, while two induced in-frame insertions and one, an in-frame deletion. The number of variants per affected infant ranged from one to six, with a mean number of 2.5 variants per infant. Four variants were recurrent. Two variants in *CALM2* occurred in 3 infants, one was homozygous for one of these *CALM2* variants and two infants were heterozygous for each *CALM2* variant. One *CFTR* and one *ATM* variant each occurred in two infants as well. For individual genes, the number of variants ranged from 1 to 13. Fifty-six infants were found with only one of these variants of interest, however, multiple variants were identified in several decedents. Twenty-two infants were found with two variants. Four infants were found with three variants, and five infants were found with four variants. Three infants had five of these variants, and two were found with six variants. Forty-two of these genes can be characterized as cardiac genes, 23 as neurologic, while 13 are related to systemic function. An additional eight genes are related to various syndromes and are found in the Reactive Oxygen Species (ROS) pathway.

### 4.2. Reactive Oxygen Species (ROS) Pathway Related Genes (Table 2)

The previously reported pathogenic variants included 20 in eight genes found in the antioxidant pathway that plays a critical role in the hypoxic response, the ROS pathway, *AAGAB*, *ATM*, *BRCA1*, *CFTR*, *COL7A1*, *ITGB3*, *LAMB3*, and *SMAD3*. (Figure 7). ROS pathway variants were identified in 21 infants (13.8%). One infant had two of these variants and two of the same variants were found in two infants each, as previously stated. Exceptions to the inclusion of ultra-rare variants include two *CFTR* variants, and one *ITBG3* variant. These variants were found in infants of African (A) descent and are more common in that population, not meeting our criteria for rare/ultra-rare. However, these variants are reported here due to the overall rarity of the allele in the combined total population and the previously reported pathogenicity in ClinVar. See Table 2.

**Figure 7.**
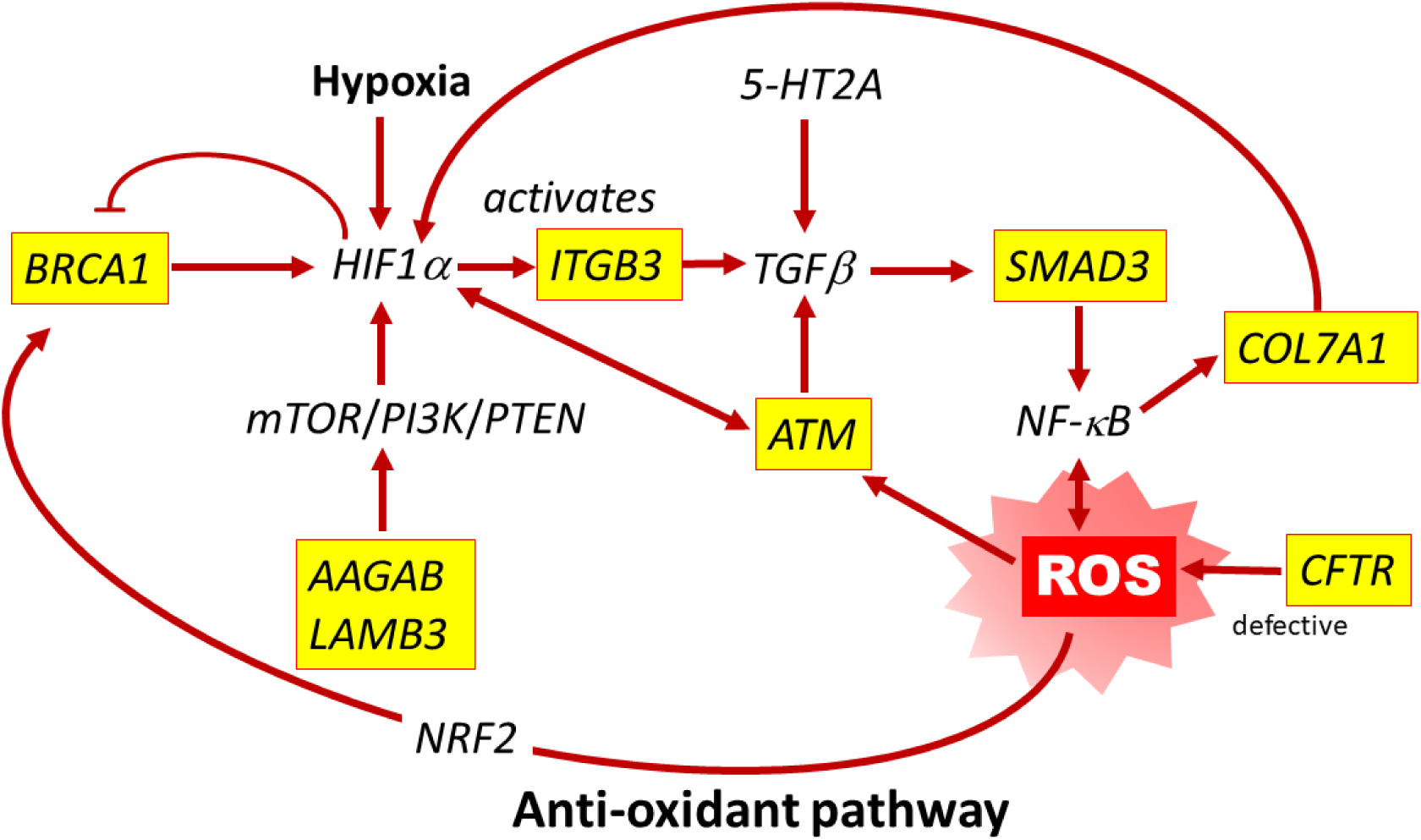
ROS Pathway. Twenty variants in eight genes (*AAGAB, ATM, BRCA1, CFTR, COL7A1, ITGB3, LAMB3 and SMAD3*) that function in the ROS pathway were found in 21 infants who succumbed to SUID.

**Table 1.** Cohort demographics as reported by parents, death scene investigators, Medical Examiners.

| Demographics | Number of Cases | Proportion |
| --- | --- | --- |
| Ethnicity/Race |  |  |
| White | 73 | 50.3% |
| Black or African American | 63 | 43.4% |
| Hispanic | 3 | 2.1% |
| American Indian | 2 | 1.4% |
| Not specified | 4 | 2.8% |
| Sex |  |  |
| Male | 79 | 54.5% |
| Female | 66 | 45.5% |
| Age at Death |  |  |
| < 60 days | 40 | 27.6% |
| 60 – 180 days | 93 | 64.1% |
| 181 – 365 days | 12 | 8.3% |
| Multiple Birth |  |  |
| Singlet | 134 | 92.4% |
| Twin | 10 | 6.9% |
| Triplet | 1 | 0.7% |

**Table 2.**
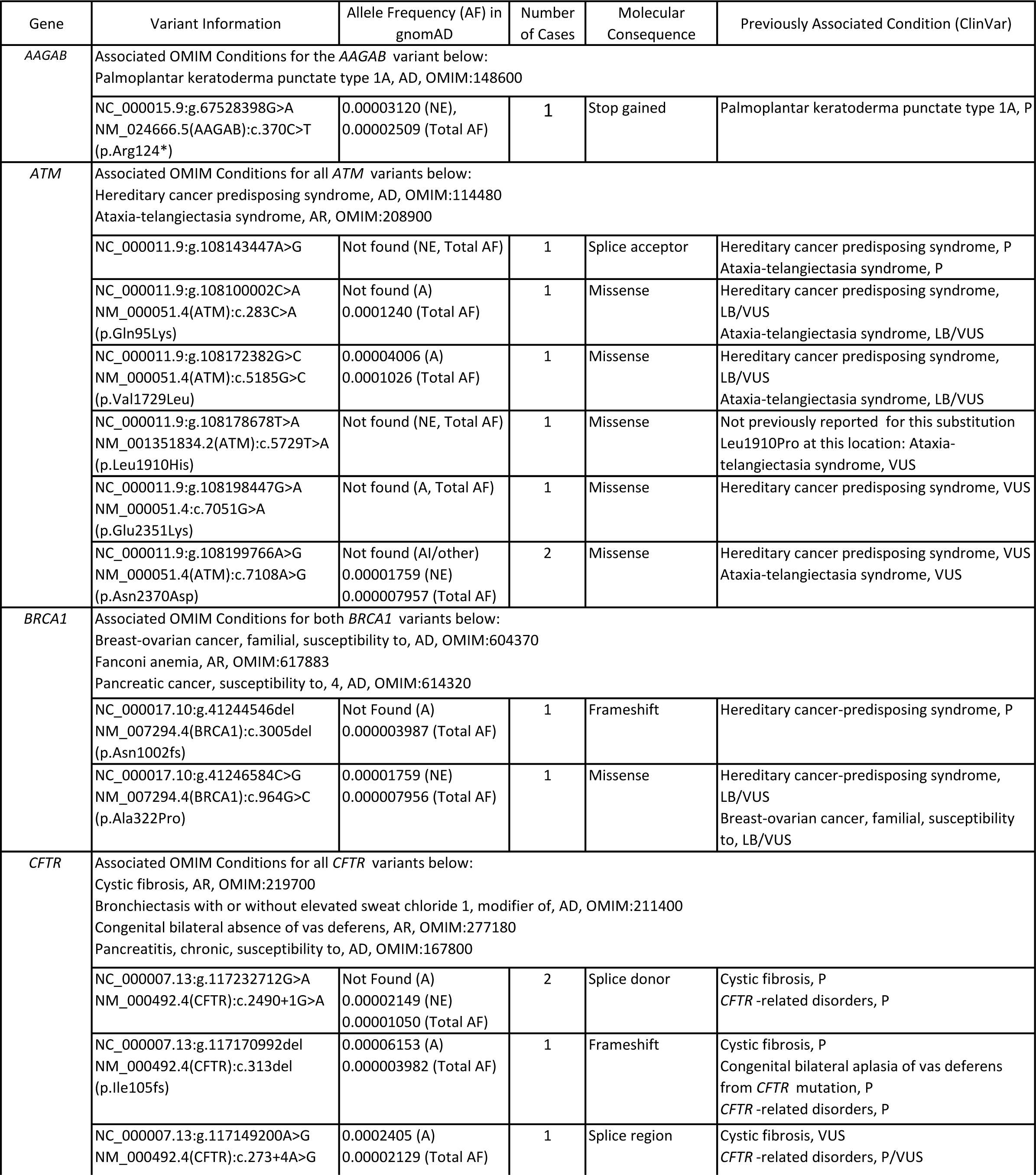
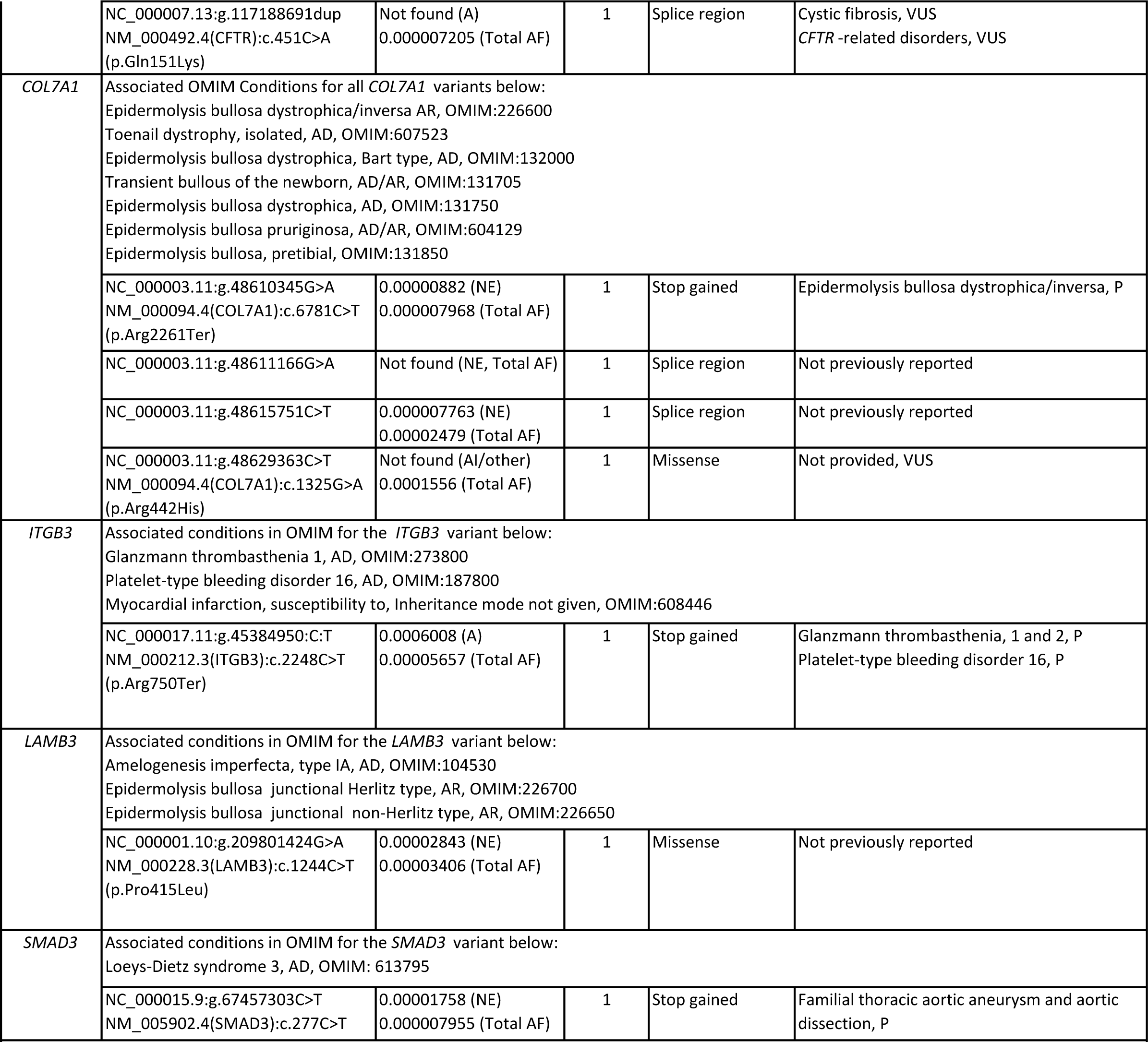
Variants in genes involved in the ROS pathway and their associated conditions in ClinVar and OMIM. A=African/African American; AD=Autosomal Dominant; AF=Allele Frequency; AI=American Indian; AR=Autosomal Recessive; B=Benign; Hisp=Hispanic; LB=Likely Benign; LP=Likely Pathogenic; NE=Northern European; P=Pathogenic; VUS = Variant of Uncertain Significance;

### 4.3. Genes previously implicated in SUID (Table 3)

In addition to the ROS pathway variants, we detected 128 variants of interest in genes that were previously implicated in SIDS/SUID/SUDP (Baruteau, 2017; Männikkö, 2018; Tester, 2018a; Tester, 2018b; Gray, 2019; Chahal, 2020; Clemens, 2020; Moore, 2020; Rochtus, 2020; Chahal, 2021; Halvorsen, 2021; Keywan, 2021; Koh, 2022). Eighty-eight of these variants occurred in 43 genes related to cardiac conditions including generalized cardiomyopathy, specific cardiomyopathies such as arrhythmogenic right ventricular dysplasia and left ventricular noncompaction; cardiac arrhythmias including atrial and ventricular fibrillation; the channelopathies, Brugada syndrome, Long QT syndrome and Catecholaminergic Polymorphic Ventricular Tachycardia (CPVT), in addition to several syndromes with a cardiac phenotype. Twenty-nine occurred in 22 genes related to neurologic function, including epilepsy, encephalopathy, ataxia, neuropathy and episodic pain syndrome, movement disorders such as dystonia and paralysis, intellectual disability and the polymalformation condition, Liang-Wang syndrome. Eleven additional variants were found in six genes related to systemic conditions and syndromes previously described in SIDS/SUID (Gray, 2019; Koh, 2022). See Table 3.

**Table 3.**
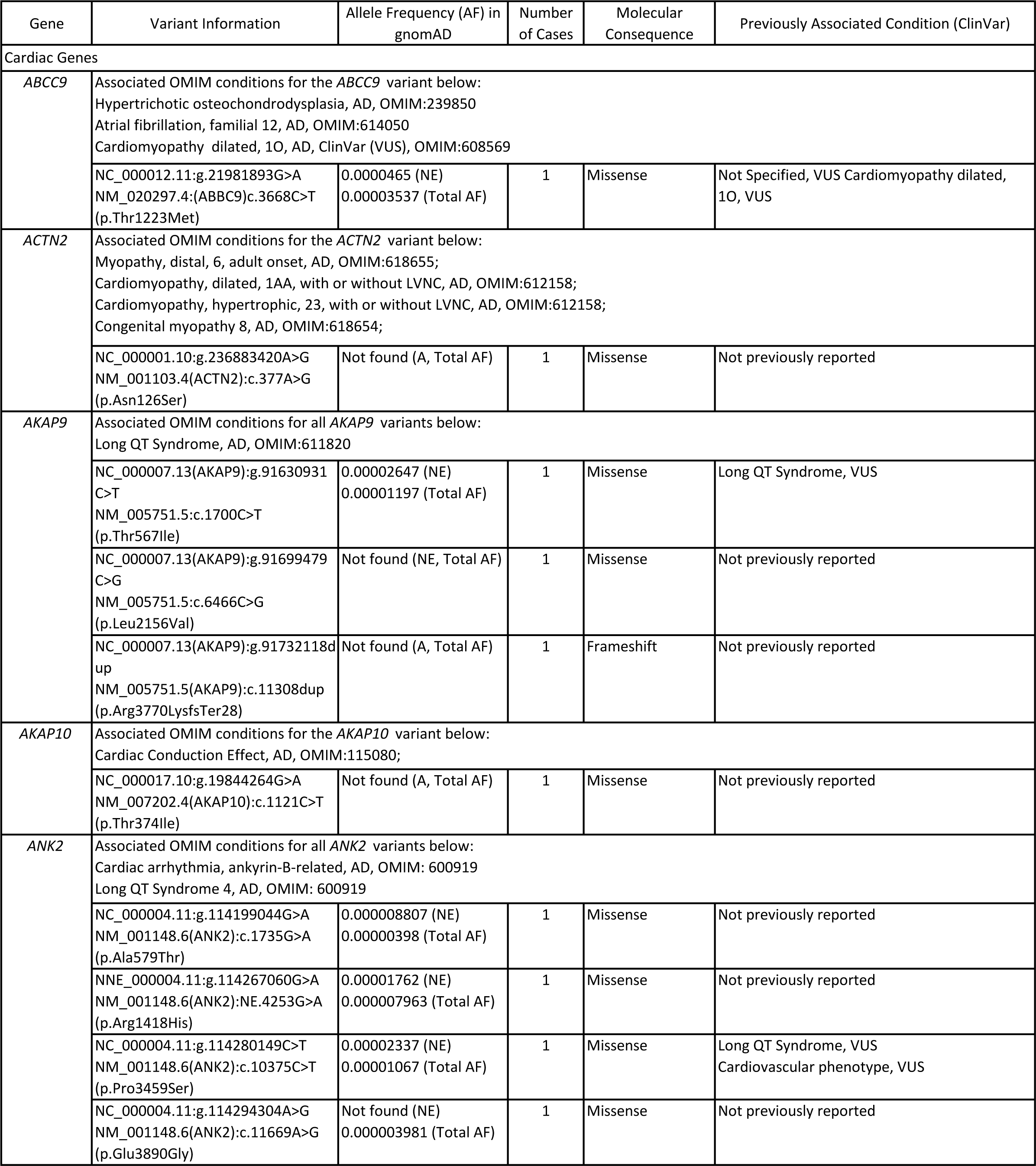
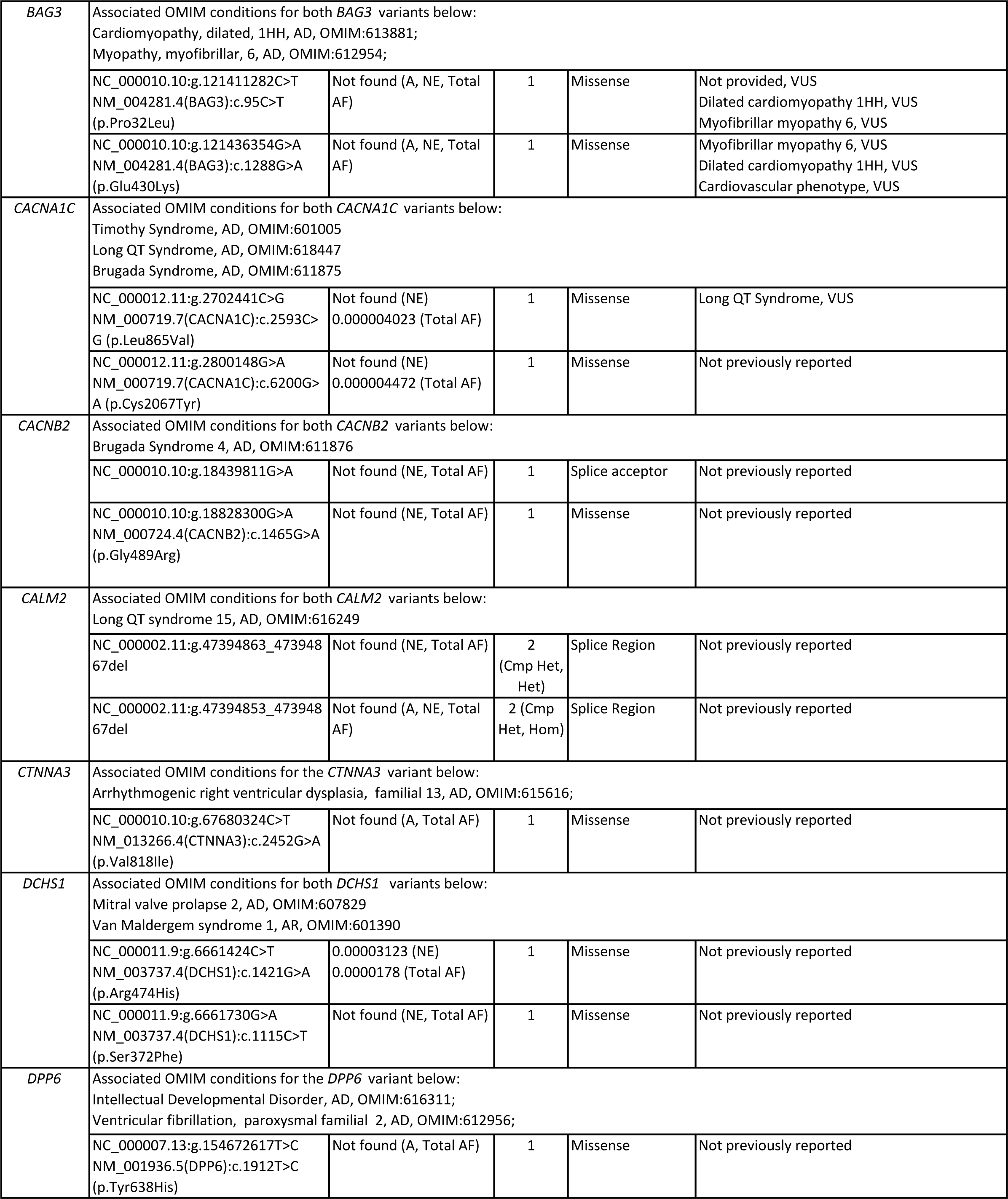
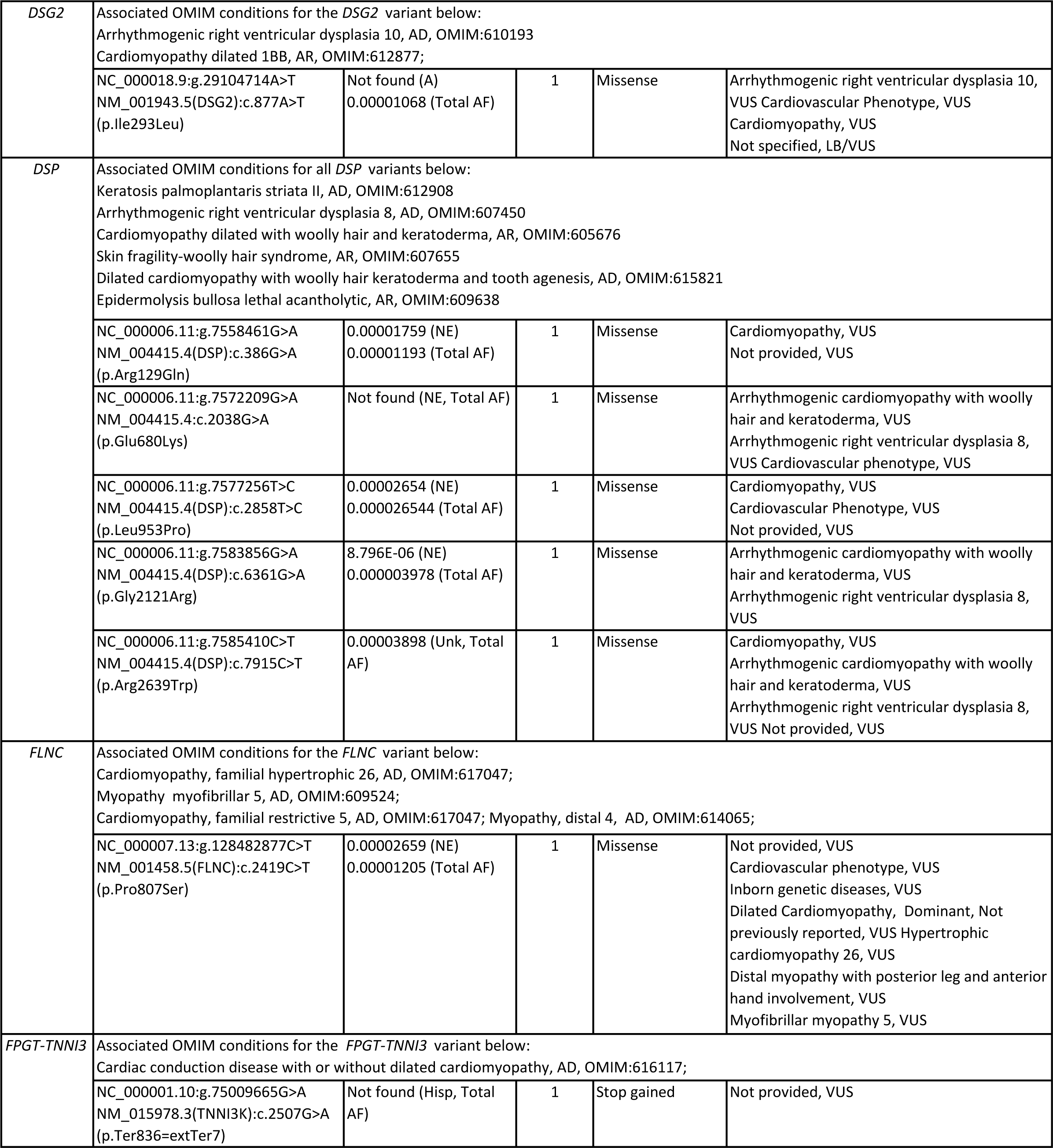
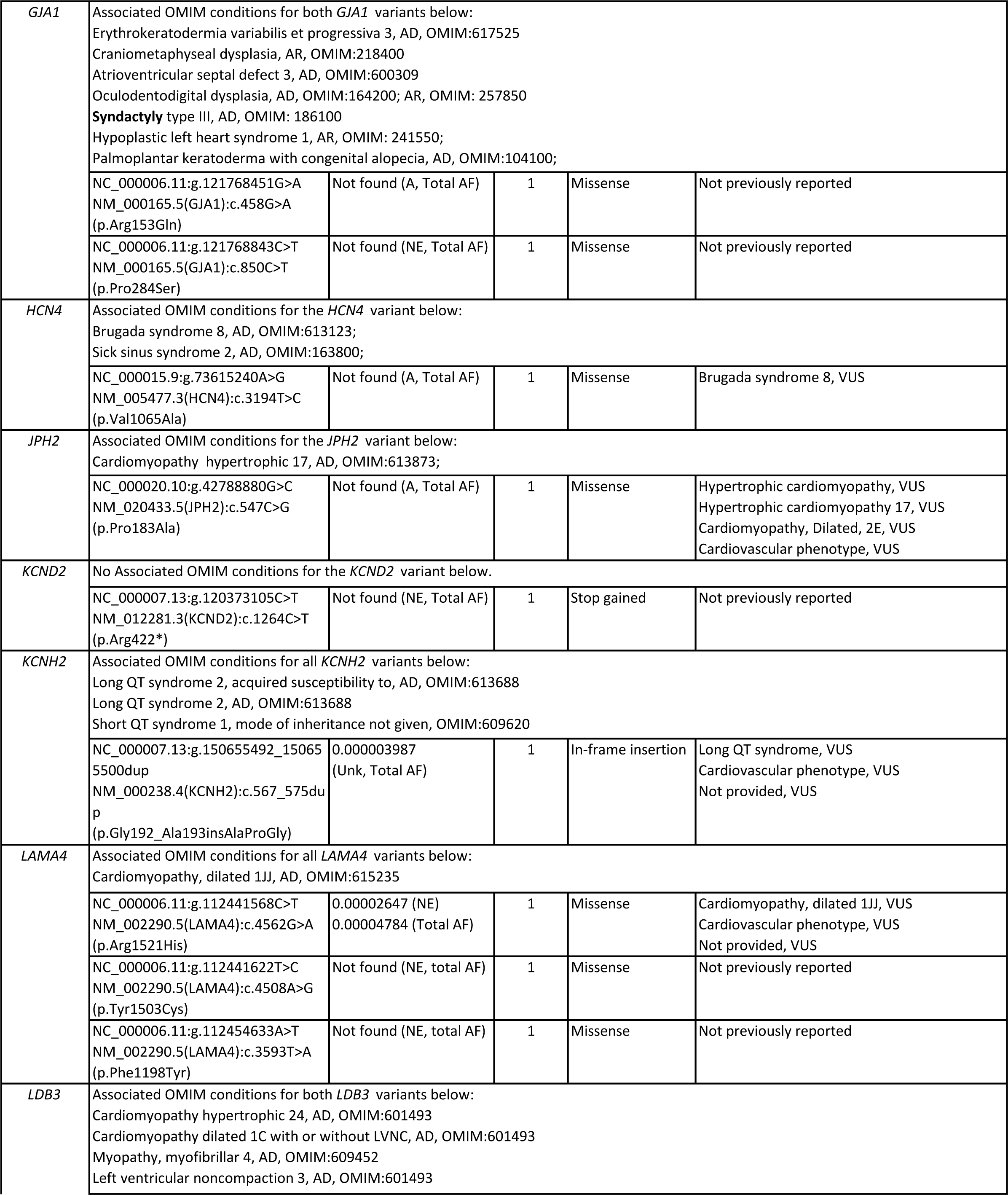
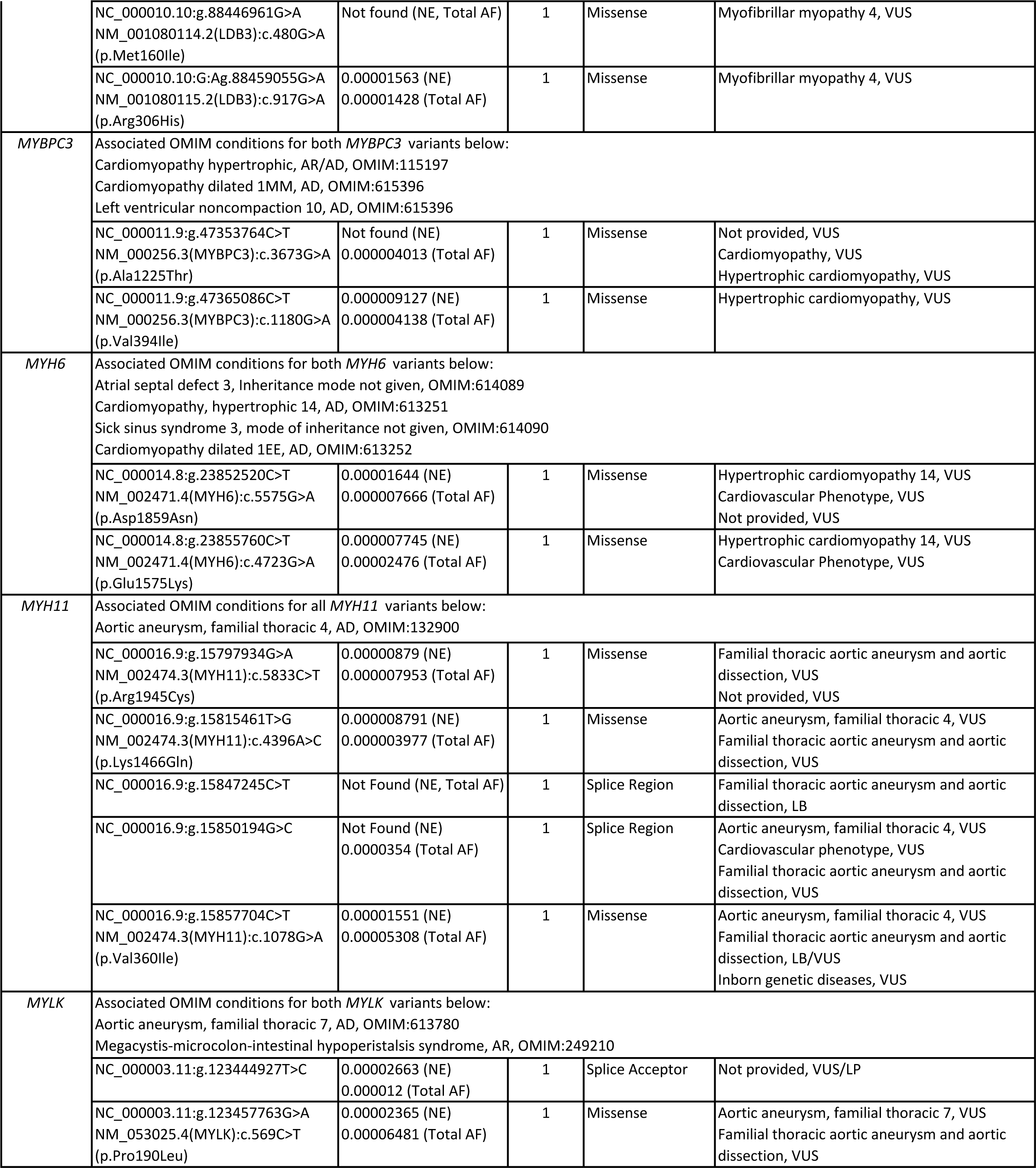
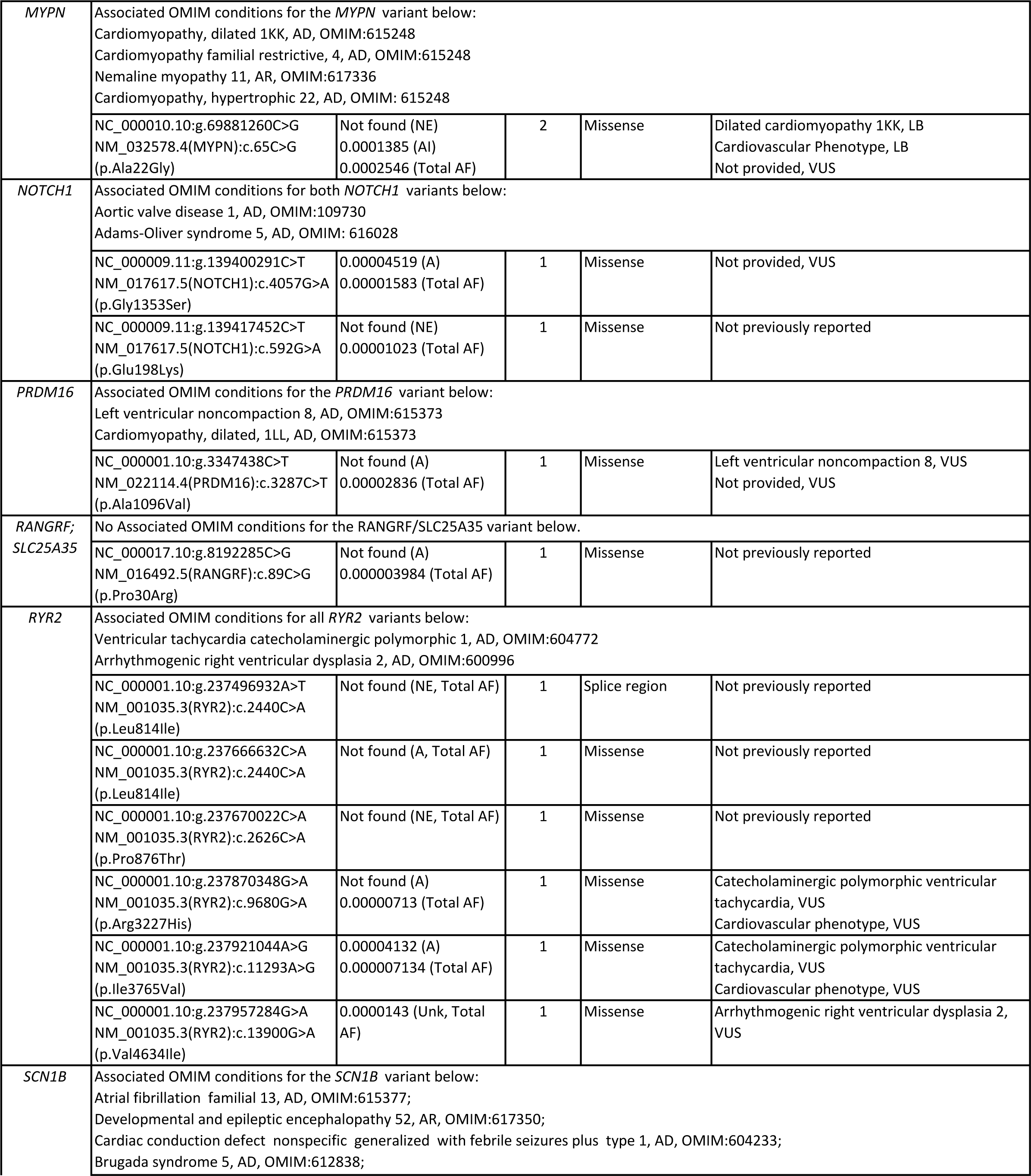
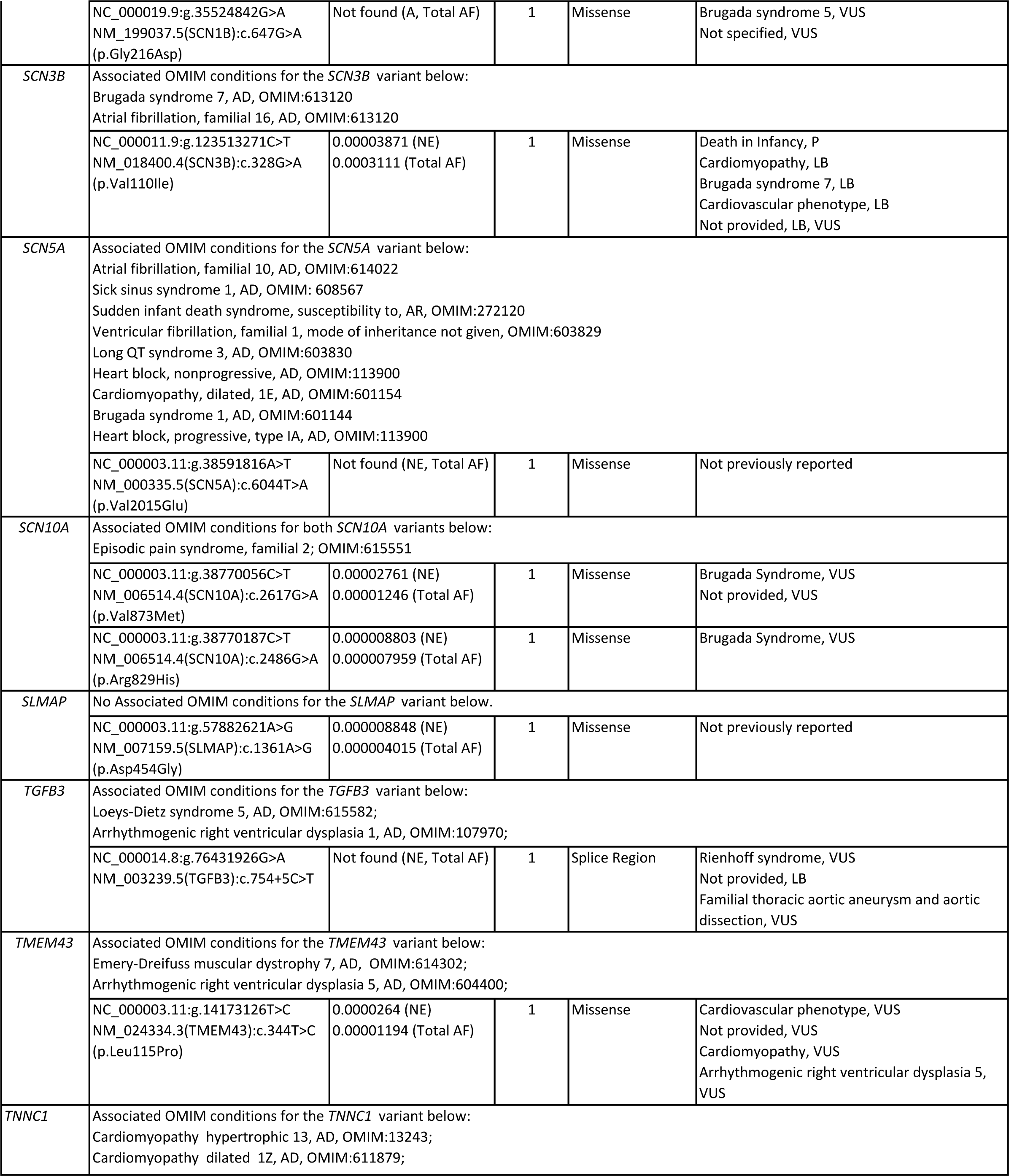
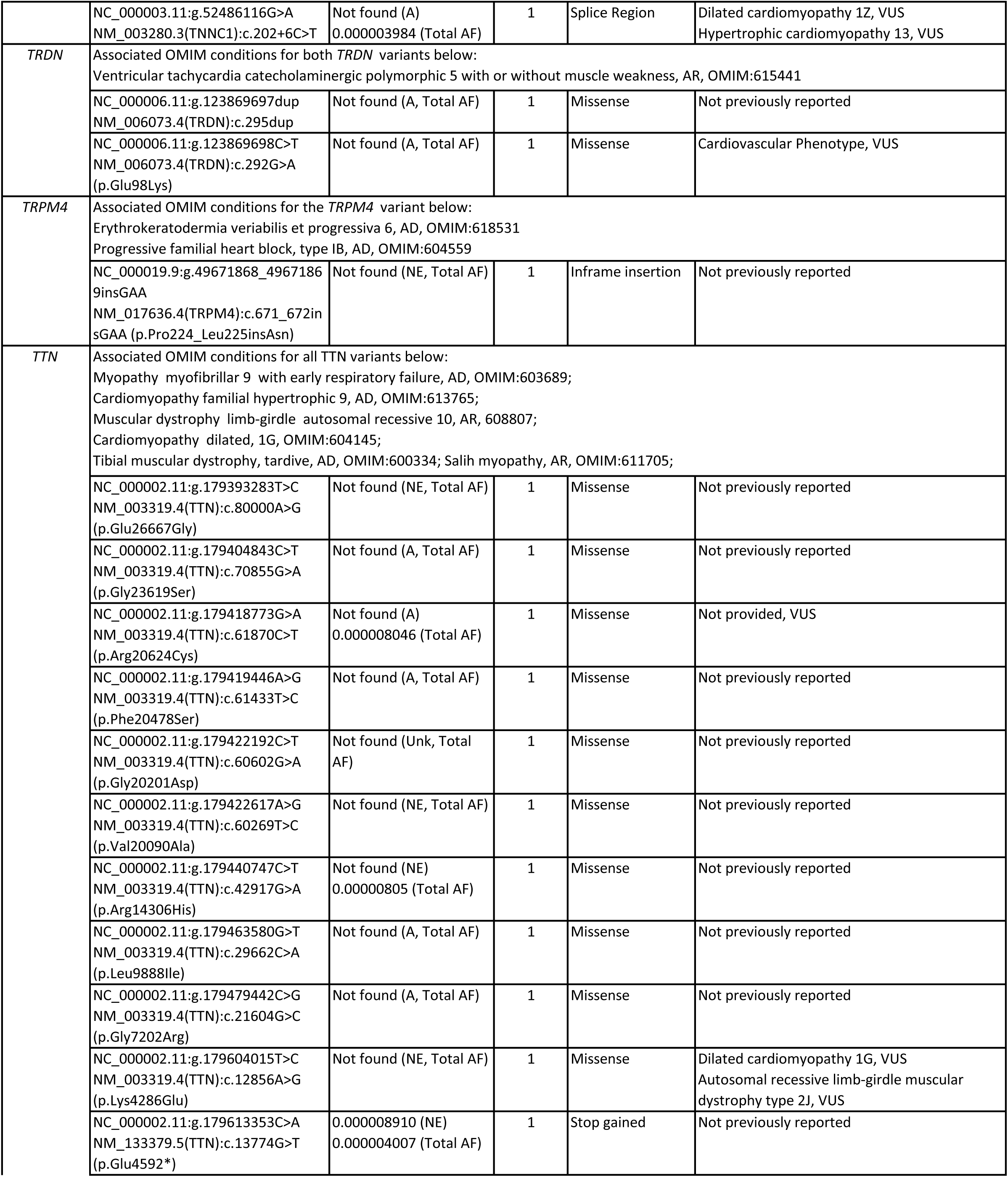
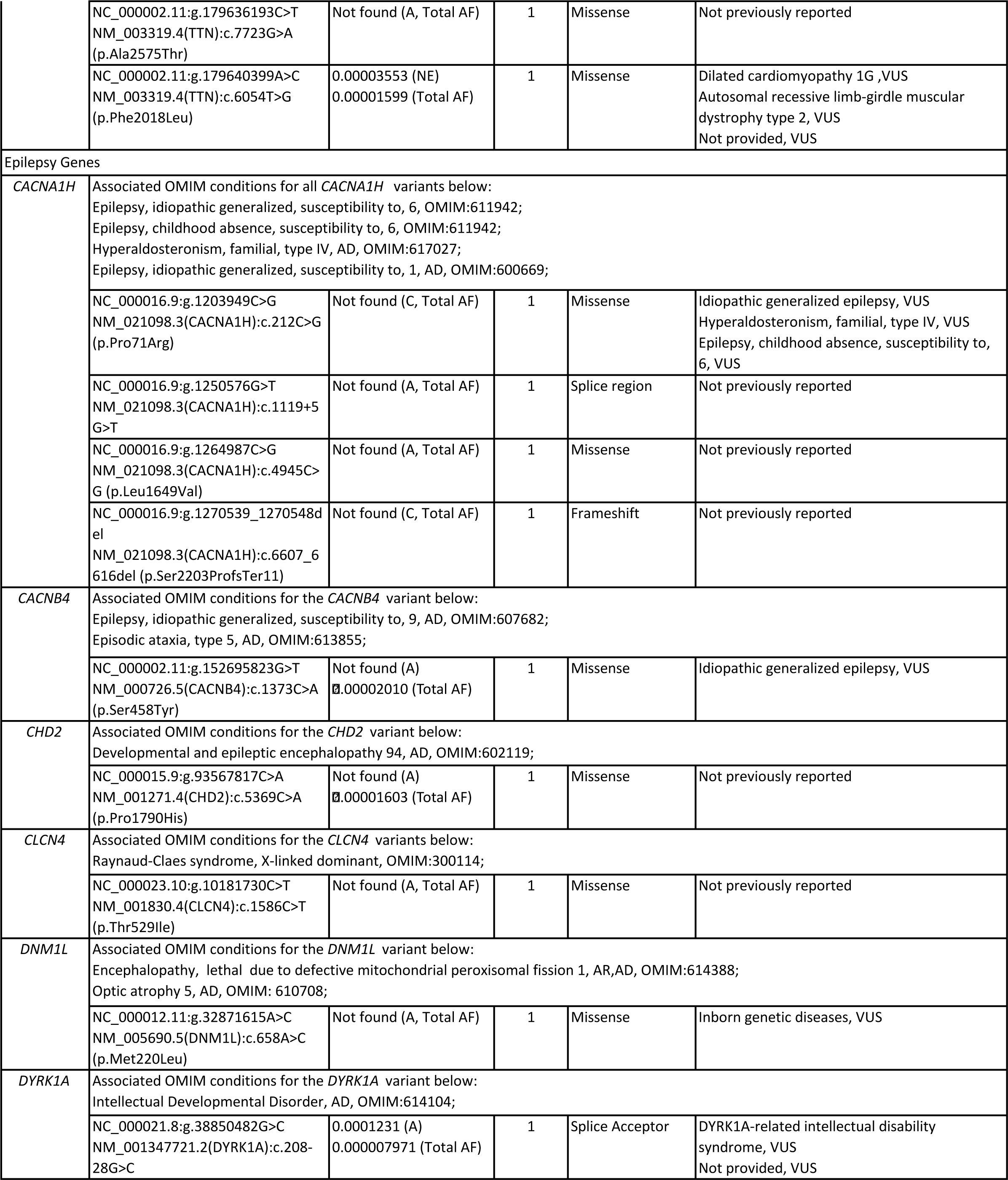
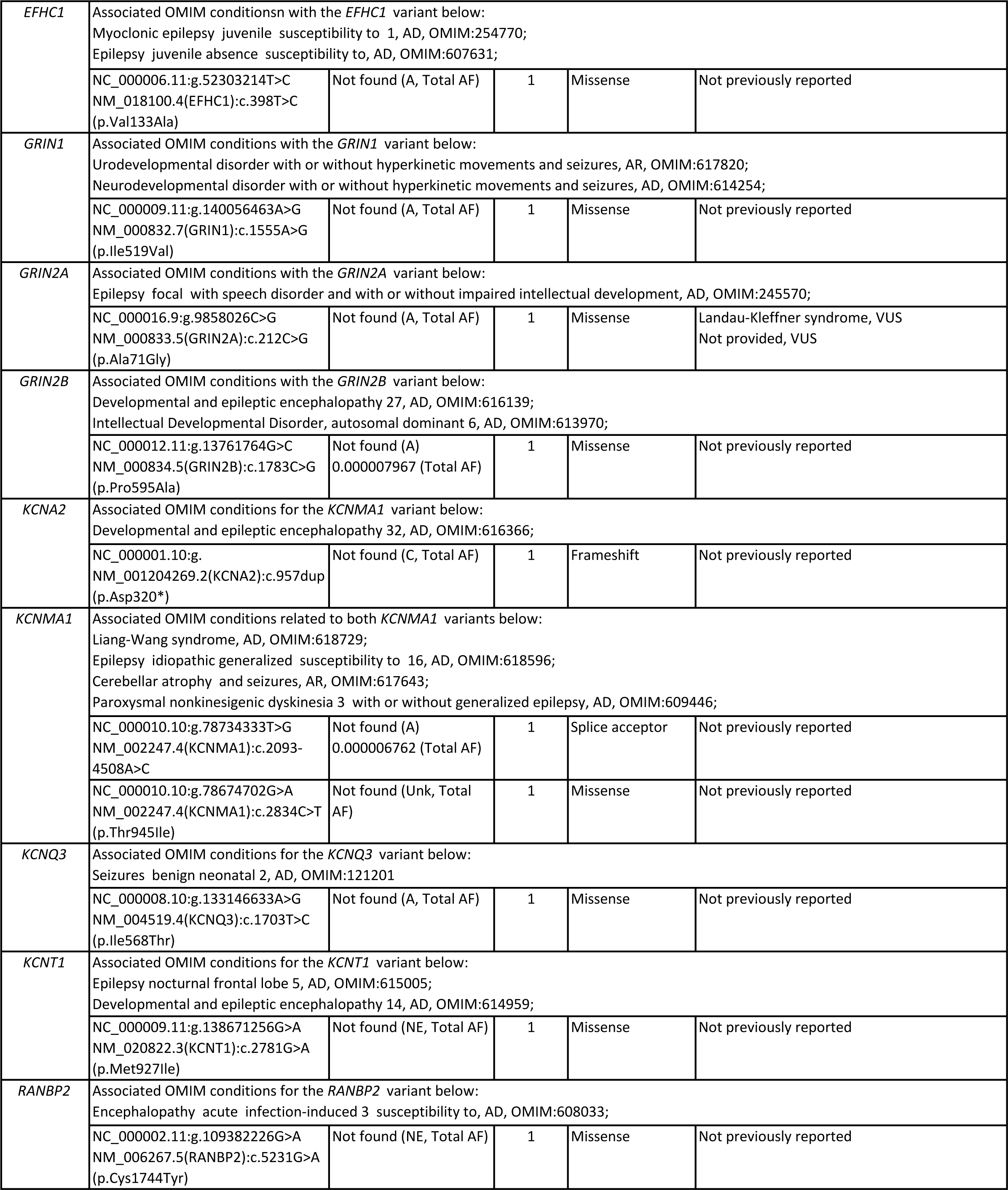
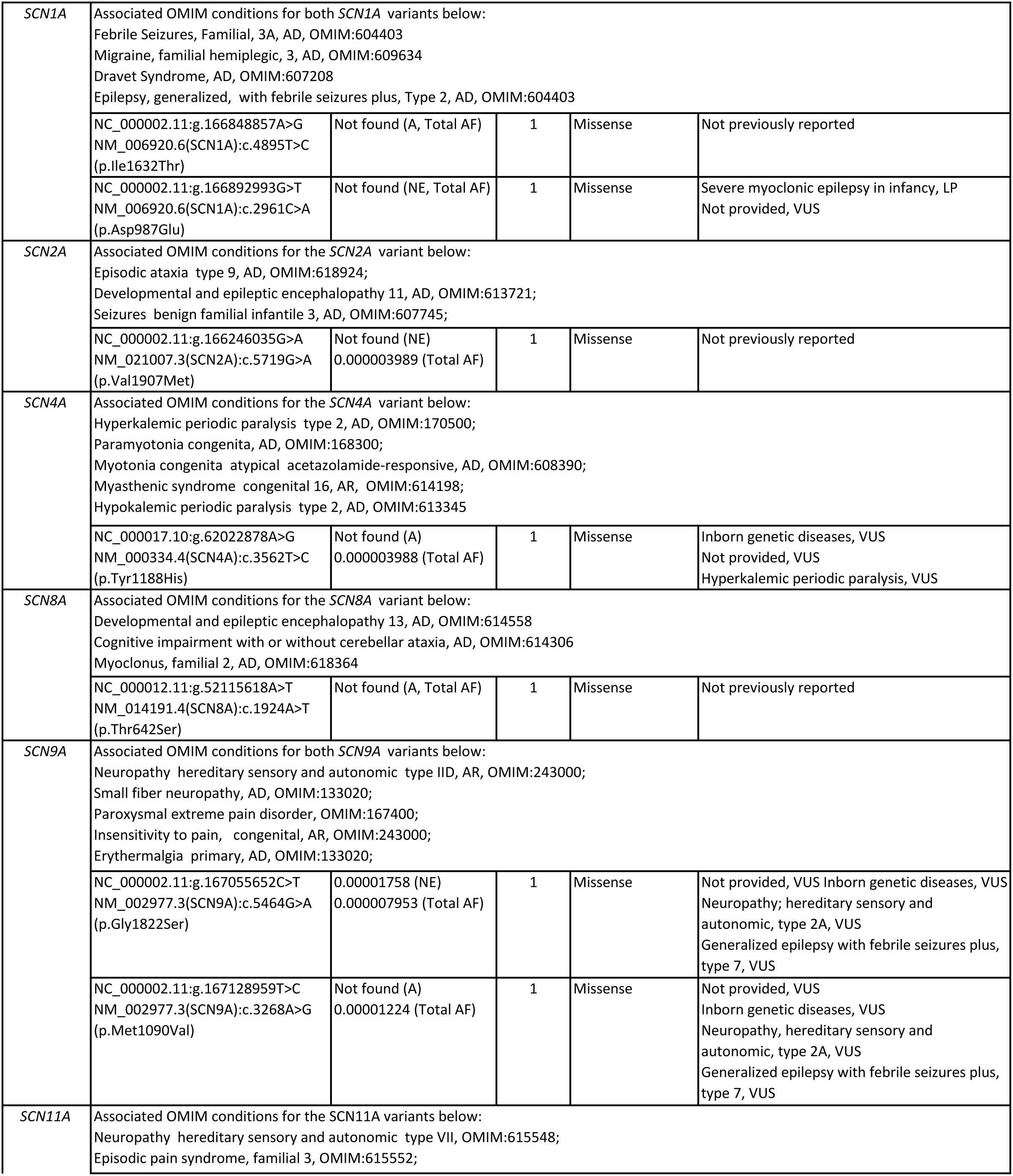
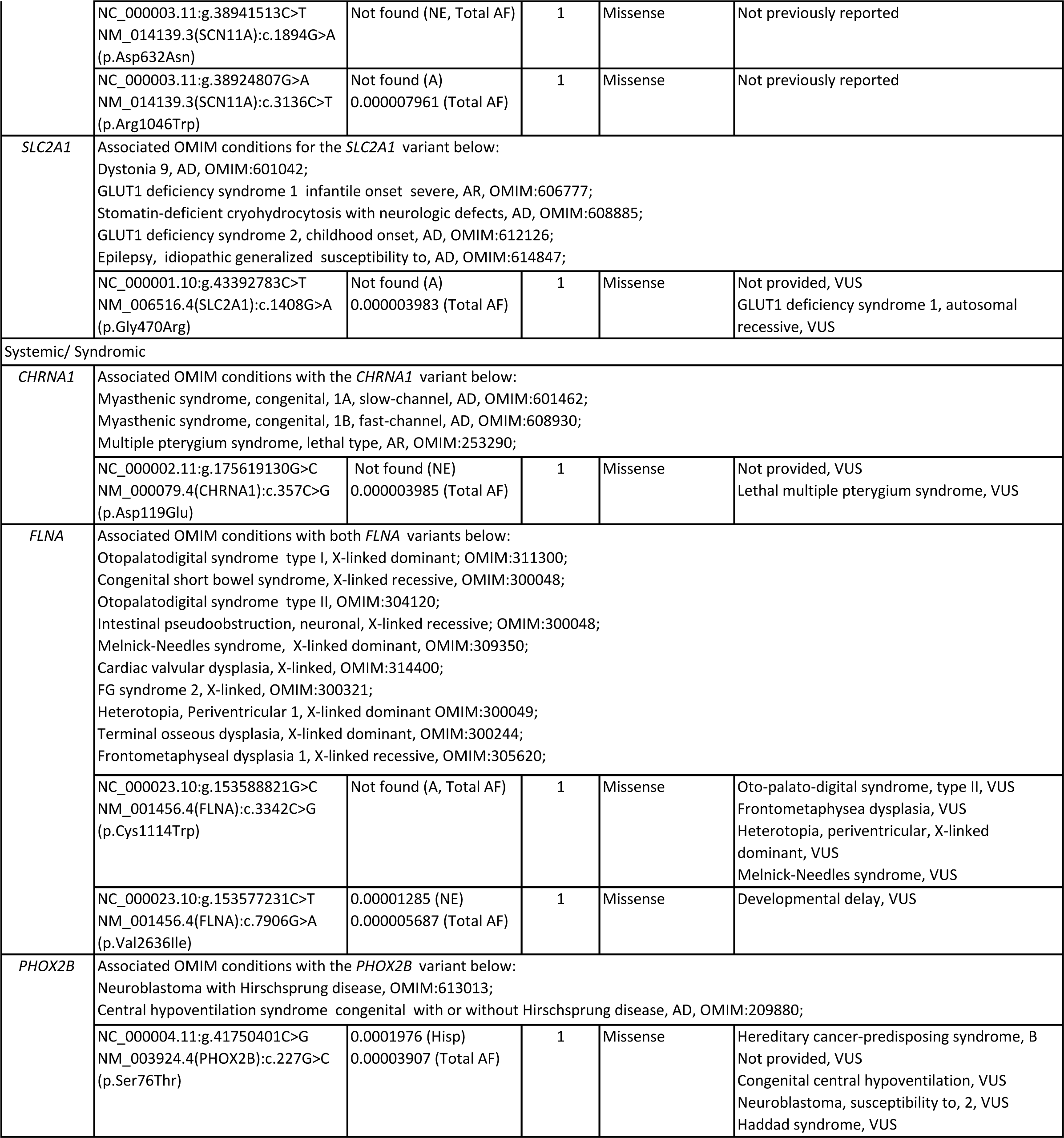
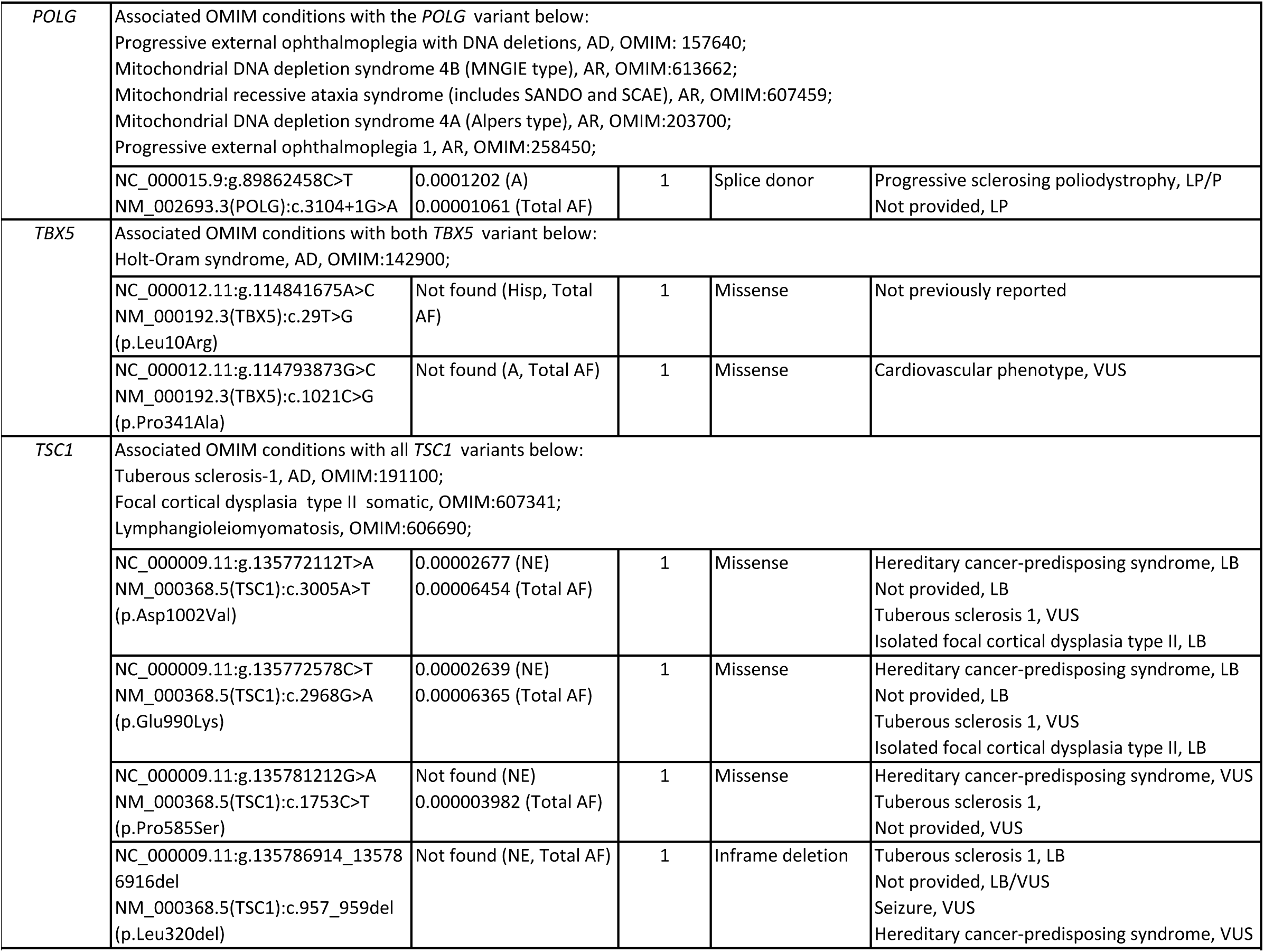
Variants in genes previously implicated in SIDS/SUID and their associated conditions in ClinVar and OMIM. A=African/African American; AD=Autosomal Dominant; AF=Allele Frequency; AI=American Indian; AR=Autosomal Recessive; B=Benign; Hisp=Hispanic; LB=Likely Benign; LP=Likely Pathogenic; NE=Northern European; P=Pathogenic; VUS = Variant of Uncertain Significance;

**Table 4.**
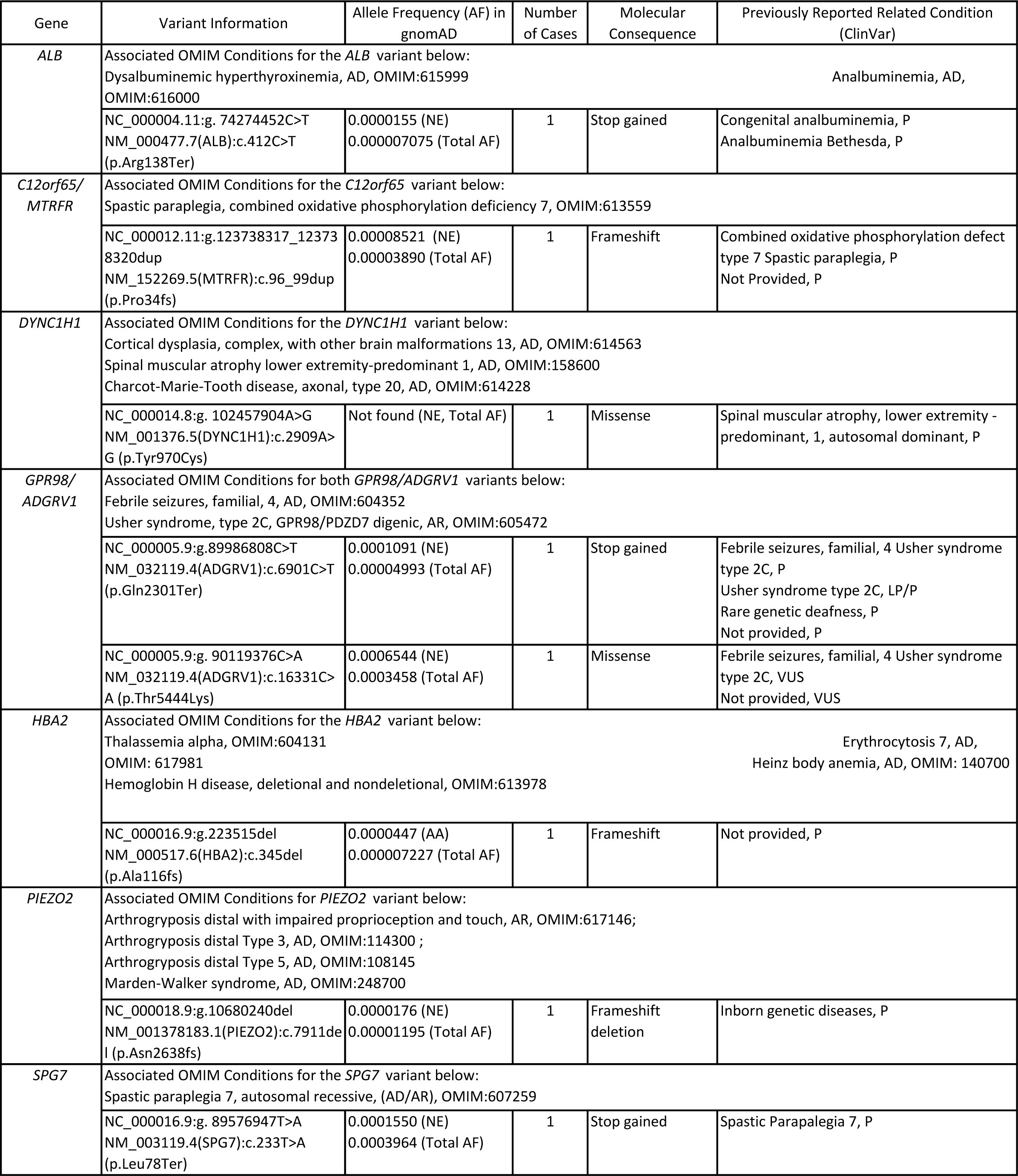
Variants in genes previously determined to be pathogenic/likely pathogenic and their associated conditions in ClinVar and OMIM. A=African/African American; AD=Autosomal Dominant; AF=Allele Frequency; AI=American Indian; AR=Autosomal Recessive; B=Benign; Hisp=Hispanic; LB=Likely Benign; LP=Likely Pathogenic; NE=Northern European; P=Pathogenic; VUS = Variant of Uncertain Significance;

#### 4.3.1. Cardiac: Cardiomyopathy

Thirty-nine variants in 17 genes associated with cardiomyopathies and related conditions were found in 37 infants – five infants had two of these variants. These include *ABCC9, ACTN2, BAG3, DSG2, DSP, FLNC, JPH2, LAMA4, LDB3, MYBPC3, MYH6, MYPN, PRDM16, SCN5A, TNNC1, FPGT-TNNI3K*, *and TTN*. Five variants in three of these genes, *JPH2, LAMA4 and TNNC1*, were found in five infants and are associated with various cardiomyopathies in the absence of other conditions (OMIM:613873, OMIM:615235, OMIM:13243, OMIM:611879). One *LAMA4* variant, Arg1521His, has been previously reported in ClinVar as a variant of unknown significance (VUS) for cardiomyopathy and an unspecified cardiovascular phenotype (ClinVar:213607). The infant that was found with this variant had a family history of SIDS. Eighteen variants in five genes associated with cardiomyopathy and myopathy, *ACTN2*, *BAG3*, *FLNC, MYPN* and *TTN,* were found in 17 infants (OMIM:613881, OMIM:612954, OMIM:617047, OMIM:614065, OMIM:617336, OMIM:615248, OMIM:618655, OMIM:612158, OMIM:618654). Five variants in three genes, *LDB3*, *MYBPC3*, and *PRDM16,* associated with cardiomyopathy and left ventricular noncompaction were identified in four infants (OMIM:601493, OMIM:609452, OMIM:115197, OMIM:615396, OMIM:615373). One infant had two of these variants. Six variants in two genes, *DSG2* and *DSP* were identified in six different children that are associated with cardiomyopathy and right ventricular dysplasia (OMIM:610193, OMIM:612877, OMIM:607450, OMIM:605676). Two variants in two infants were found in *ABCC9* and *SCN5A* that are associated with both cardiomyopathy and atrial fibrillation (OMIM:608569, OMIM:614050, OMIM:601154, OMIM:614022). The variant in *SCN5A* is also associated with ventricular fibrillation (OMIM:603829), Brugada (OMIM:601144), Long QT syndrome (OMIM:603830), heart blocks (OMIM:113900, OMIM:113900), Sick Sinus syndrome (OMIM:608567), and susceptibility to SIDS (OMIM:272120). One variant in one gene associated with cardiomyopathy and a cardiac conduction disorder, *FPGT-TNNI3K*, was found in one infant (OMIM:616117). Two *MYH6* variants were found in two different infants that were related to cardiomyopathy (OMIM:613251, OMIM:613252), Sick Sinus syndrome (OMIM:614090), and cardiac defects including atrial septal defect (OMIM:614089). One infant in our cohort was found to have atrial septal defect at autopsy and another an unspecified heart defect at autopsy, but these variants did not occur in either of these infants. There were no autopsy reports available for the infants that carried these *MYH6* variants.

#### 4.3.2. Cardiac: Congenital Conditions

Thirteen variants in five genes, *DCHS1*, *GJA1*, *NOTCH1*, *MYH11*, and *MYLK*, were found in 11 infants related to heart defects and congenital conditions in the absence of cardiomyopathy. Six variants were found in five infants in three genes associated with heart defects, *DCHS1*, *GJA1* and *NOTCH1*. These included mitral valve prolapse (OMIM:607829), atrioventricular septal defect (OMIM:600309), and aortic valve disease (OMIM:109730). One infant had a variant in both *GJA1* and *NOTCH1*. An autopsy report was available for only one of these five infants and no heart defect was found. Five *MYH11* variants associated with familial thoracic aortic aneurysm were found in four infants, one infant had two of these *MYH11* variants (OMIM:132900). Two *MYLK* variants, were found in two infants and are associated with familial thoracic aortic aneurysm (OMIM:613780). One missense (Pro190Leu) variant is reported to be a VUS for aortic aneurysm in ClinVar (ClinVar:520017). The child with the Pro190Leu variant had a family history of SUID, there were no relevant findings reported at autopsy other than the presence of focal intimal thickening in the right coronary artery.

#### 4.3.3. Cardiac: Arrhythmias and Conduction Disorders

Five variants were found in five genes associated with cardiac arrhythmias and conduction disorders alone such as cardiac conduction effect, *AKAP10* (OMIM:115080), arrhythmogenic right ventricular dysplasia, *CTNNA3* (OMIM:615616), ventricular fibrillation*, DPP6* (OMIM:612956), arrhythmogenic right ventricular dysplasia, *TGFB3* (OMIM:107970), and *TMEM43* (OMIM:604400). These variants were found in five infants.

#### 4.3.4. Channelopathies: Brugada syndrome, Long QT syndrome and CPVT

Thirty-two variants were found in 27 decedents in 17 genes related to the channelopathies Brugada and Long QT syndromes and CPTV. Nine variants in seven genes, *CACNB2, HCN4, KCND2, RANGRF, SCN10A, SLMAP, and TRPM4* were identified in eight decedents that have been implicated in Brugada syndrome in previous studies (Baruteau, 2017; Tester, 2018b; Rochtus, 2020; Keywan, 2021; Koh, 2022). Two of these variants in genes related to Brugada syndrome, *SCN10A* and *SLMAP,* were found in one infant. The *SCN10A* variant has been previously reported in ClinVar as a VUS for Brugada syndrome (ClinVar:641909) and also associated with the neurologic condition, episodic pain syndrome (OMIM:615551). The *SLMAP* variant has not been previously reported. Three variants in *SCN1B*, *SCN3B*, and *SCN5A* found in three infants, have been previously associated with Brugada syndrome as well as other arrhythmias (OMIM:612838, OMIM:613120, OMIM:601144). The *SCN3B* variant found has also been previously reported as P for death in infancy (ClinVar:213607). The *SCN5A* variant as previously discussed in section 4.3.1, has been associated with Long QT and SIDS among other conditions. The mother of the child in which the *SCN1B* variant was found was noted to have lost another child at three months of age to SIDS. Two infants were found with two additional variants in one gene, *CACNA1C*, that has been associated with Brugada syndrome (Chahal, 2020; Halvorsen, 2021; Koh, 2022; OMIM:611875) in addition to other arrhythmias (OMIM:601005, OMIM:609620), and Long QT syndrome (OMIM:618447). Six variants were found in three genes that have previously been linked to Long and/or Short QT Syndrome alone, *AKAP9*, *CALM2* and *KCNH2* (Baruteau, 2017; Tester, 2018a; Tester, 2018b; Chahal, 2020; Clemens, 2020; Moore, 2020; Keywan, 2021; Koh, 2022; OMIM:611820, OMIM:616249, OMIM:613688, OMIM:609620). A few children were found to have more than one of these variants. One infant had one variant in both *CACNA1C* and *AKAP9*, and one infant had two different *CALM2* variants, while a third infant was homozygous for one of these *CALM2* variants. Four variants in *ANK2* were found in four infants. Variants in this gene have been linked to Long QT syndrome in addition to other arrhythmias previously (Baruteau, 2017; Chahal, 2020; Koh, 2022; OMIM:600919). The infant that was found with 2 different *CALM2* variants also had one of these *ANK2* variants. Six variants in *RYR2* were found in six infants. Variants in *RYR2* have been associated with CPVT (Baruteau, 2017; Tester, 2018b; Chahal, 2020, 2021; Halvorsen, 2021; Koh, 2022, OMIM:604772) and arrhythmogenic right ventricular dysplasia (OMIM:600996). One infant that had a variant in *RYR2* was also the infant found to be homozygous for one of the *CALM2* variants. Two variants in *TRDN* were identified in one infant. Variants in *TRDN* have been associated with CPVT through an AR mode of inheritance in OMIM (OMIM:615441).

### 4.4. Neurologic Disorders

Twenty-nine variants were found in 22 genes in 26 infants previously implicated in SIDS/SUID/SUDP that are related to neurologic disorders. Three infants had two of these variants each. Eleven variants in eleven infants were found in seven genes related to epilepsy, four in *CACNA1H*, one each in *CACNB4*, *EFHC1*, *GRIN1*, *GRIN2A*, *KCNQ3* and two in *SCN1A* previously implicated in SIDS/SUID/SUDP (Baruteau, 2017; Tester, 2018; Rochtus, 2020; Keywan, 2021; Koh, 2022; OMIM:611942, OMIM:600669, OMIM:607682, OMIM:254770, OMIM:607631, OMIM:614254, OMIM:245570, OMIM:121201, OMIM:604403, and OMIM:607208). One *SCN1A* missense variant, Asp987Glu, has been previously reported in ClinVar to be LP for severe myoclonic epilepsy in infancy (SMEI), also known as Dravet syndrome (ClinVar:520017), a condition which has been previously implicated in SIDS/SUID/SUDP and SUDEP (Gray, 2019; Chahal, 2020; Rochtus, 2020; Chahal, 2021; Keywan, 2021; Koh, 2022). Five variants were found in four genes in five children related to epilepsy and other conditions *KCNMA1*, *KCNT1*, *SCN2A* and *SLC2A1* (OMIM:618596, OMIM:617643, OMIM:609446, OMIM:615005, OMIM:614959, OMIM:613721, OMIM:607745, OMIM:618924, OMIM:614847, OMIM:608885) Two infants were found with two different *KCNMA1* variants associated with the polymalformation condition, Lian-Wang syndrome (OMIM:618729). The infant with the *KCNQ3* variant also had a variant in *SLC2A1* associated with dystonia (OMIM:601042), childhood onset of GLUT1 deficiency syndrome 2 of AD inheritance (OMIM:612126), Stomatin-deficient cryohydrocytosis with neurologic defects, AD, (OMIM:608885) as well as epilepsy (OMIM:614847). This infant was noted to have a “well developed infant brain with isolated microglial nodules” by the ME. Five variants in five genes were found in five infants related to encephalopathy, *CHD2, DNM1L, GRIN2B*, *KCNA2*, and *RANBP2* (OMIM:602119, OMIM:614388, OMIM:616139, OMIM:616366, OMIM:608033). One *SCN8A* variant found in one infant was associated with both epilepsy and encephalopathy (OMIM:614558, OMIM:618364). The infant with the *RANBP2* variant also had the *KCNT1* variant, related to epilepsy (OMIM:615005), as previously stated, and encephalopathy (OMIM:614959). The infant with the *DNM1L* variant was noted at autopsy to have “neuronal changes consistent with hyperacute hypoxia ischemia”. Two additional infants were found to have one variant each in *SCN11A*, related to episodic pain syndrome and neuropathy (OMIM:615552, OMIM:615548). Another two children were found with a variant each in *SCN9A* also related to neuropathy (OMIM:243000, OMIM:133020). Two variants were identified in two children related to intellectual disability, *CLCN4* and *DYRK1A* (OMIM:300114, OMIM:614104) The infant with the *DYRK1A* variant also had the *KCNMA1* variant. Finally, one *SCN4A* variant was identified in one infant related to movement disorders and paralysis (OMIM:168300, OMIM:608390, OMIM:170500, OMIM:613345). This variant is reported as VUS in ClinVar for Hyperkalemic periodic paralysis (ClinVar:1899177).

### 4.5. Systemic/Syndromic Conditions (Tables 3 and 4)

Eleven variants were found in six genes in eight infants that are reported to be VUS, LP, or P for various syndromes for which there was no noted diagnosis at the time of death. Variants in six of these genes have been previously implicated in SIDS/SUID/SUDP, including *CHRNA1, FLNA, PHOX2B, POLG, TBX5, and TSC1* (Koh, 2022). The variant in *CHRNA1* has been linked to Lethal multiple pterygium syndrome, (ClinVar:466178). The variant in *FLNA* has been associated with multiple conditions in OMIM including, Otopalatodigital syndrome (OMIM:311300), Melnick-Needles syndrome (OMIM:309350), Cardiac valvular dysplasia (OMIM:314400), FG syndrome (OMIM:300321), Periventricular heterotopia (OMIM:300049), Terminal osseous dysplasia (OMIM:300244). The variant in *PHOX2B* has been implicated in Central hypoventilation syndrome (VUS, ClinVar:486030, OMIM:209880). The *POLG* variant has been associated with Progressive sclerosing poliodystrophy (LP, ClinVar:619308) and Progressive external ophthalmoplegia with DNA deletions (OMIM:157640). The *TBX5* variant has been previously associated with Holt-Oram syndrome (OMIM:142900). Both of the *TSC1* variants have been associated with Tuberous Sclerosis (LB/VUS, ClinVar:237715, LB/VUS, ClinVar:237714, OMIM:191100), Focal cortical dysplasia (OMIM:607341) and Lymphangioleiomyomatosis (OMIM:606690). The infant in which the *PHOX2B* variant was identified was noted to be "in the 10th percentile of growth for age”, and to have “mesenteric lymphadenopathy, hepatosplenomegaly, adrenal hypoplasia” at autopsy. This infant was also found to have one of the *TBX5* variants. The child in which the *CHRNA1* variant was found was also found to have a *DYNC1H1* variant (discussed in the next paragraph) previously reported as P in ClinVar for spinal muscular atrophy, a condition of AD inheritance (ClinVar:30034). This child was noted to have been hospitalized for 10 days following birth for unspecified reasons. See Tables 3 and S6.

Eight rare (<0.01%) variants were incidentally found in seven genes that are reported to be associated in ClinVar and OMIM with various syndromes, *ALB* (P, Congenital analbuminemia, ClinVar:156320, OMIM:616000), (Dysalbuminemic hyperthyroxinemia, OMIM:615999); *C12orf65/MTHFR* (P, Spastic paraplegia, ClinVar:214192, OMIM:613559); *DYNC1H1* (P, Spinal muscular atrophy, ClinVar:30034, OMIM:158600), *GPR98/ADGRV1* (P, Usher Syndrome, Febrile Seizures, ClinVar:6798, OMIM:604352, OMIM:605472); *HBA2* (P, Not provided, ClinVar:439116, Thalassemia alpha, OMIM:604131, Hemoglobin H disease, OMIM:613978, Heinz body anemia, OMIM: 140700); *PIEZO2* (P, Inborn genetic diseases, ClinVar:521668, Arthrogryposis distal, OMIM:114300, OMIM:108145, OMIM:617146, Marden-Walker syndrome, OMIM:248700); and *SPG7* (Spastic Parapalegia, P, ClinVar:6816, OMIM:607259); Variants in these genes have not been previously associated with SIDS/SUID. Two *GPR98/ADGRV1* variants in one infant were identified that are related to familial febrile seizures and Usher syndrome, both of digenic and AR inheritance. The first results in a premature stop and has been previously reported to be P for these conditions (ClinVar:6798). The second variant is a missense variant that is reported to be a VUS for these conditions (ClinVar:198666). This variant is found more commonly in the Northern European (NE) population (AF=0.065%, NE; Total AF=0.035%) and does not meet our criteria for rare. (The infant in which this variant was found was white.) Due to the mode of inheritance of these conditions, this variant is also likely significant for the development of disease, so both are included here. See Tables 4 and S6.

### 4.6. Group Analysis: Average Age at Death

The age of death was adjusted according to gestational age for those infants that were noted to have been born prematurely. Infants were grouped according to the type and number of variants each were found to have, cardiac, neurologic, ROS pathway, and systemic/syndromic. The average age of death was calculated for each group among children that were found to have only one variant. A single variant in a cardiac related gene was found in 31 infants for whom the age of death was known. Thirteen infants were found with a single variant related to neurologic function whose age of death was known and 12 infants were identified with a single ROS pathway gene variant. There were only two infants with one variant among the systemic/syndromic group, so this group was excluded from the analysis. The median age at death for 29 infants with a single cardiac gene variant was 10.1 weeks (IQR=8.9, 13.0), whereas for the 14 infants that were found to have a single variant in a gene related to neurologic function, the median age at death was 16.1 weeks (IQR=11, 21.3). The 11 infants with a single variant in a ROS pathway gene died at the median age of 12.7 weeks (IQR=9.4, 21.4). ANOVA was run in R and revealed that these differences were significant (P=0.019) (Figure 8). See Table S7 for a full list of abbreviations used in this manuscript.

**Figure 8.**
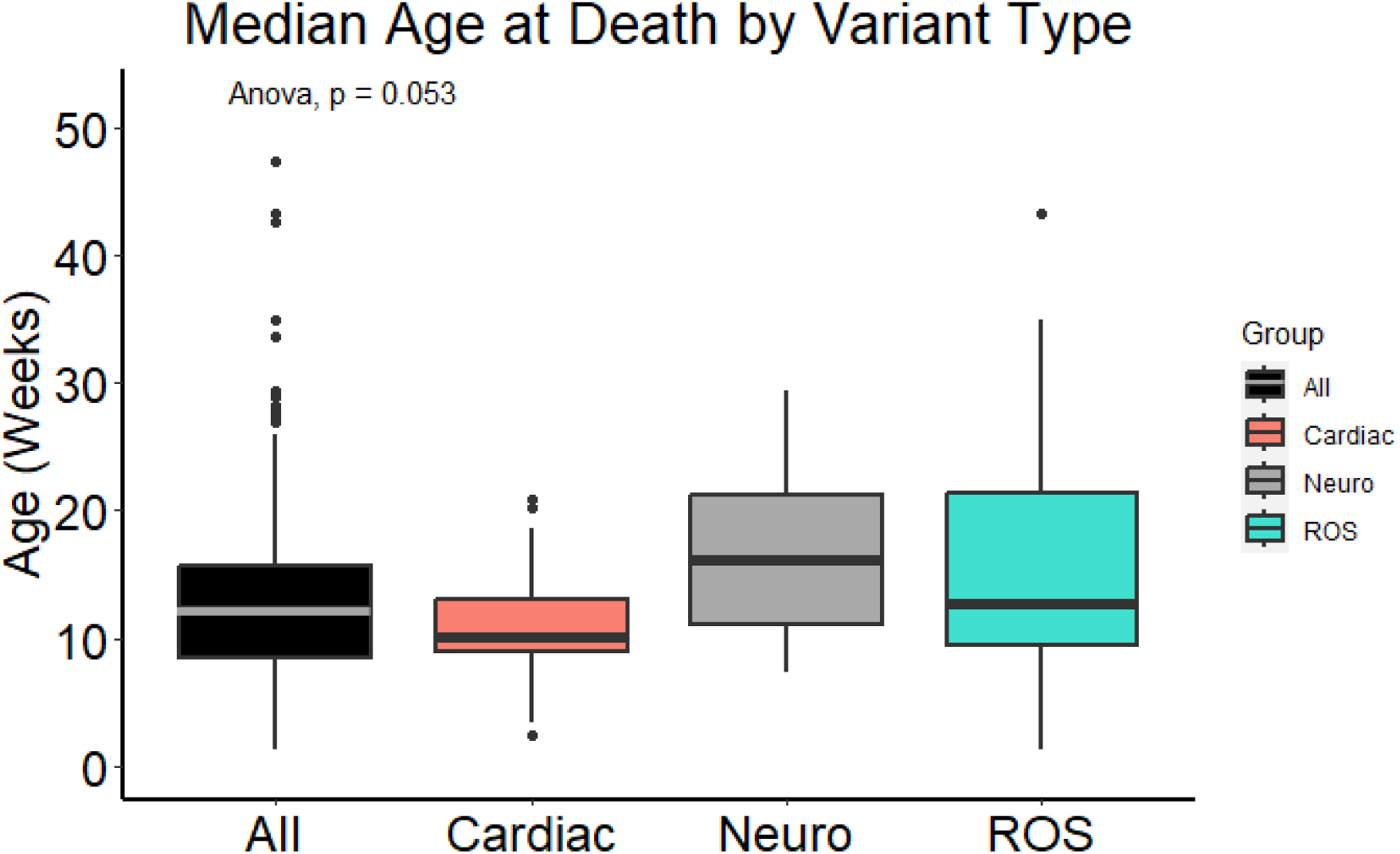
Median age at death by variant type in affected infants. Median age at death was 12.2 weeks (IQR=8.4, 15.8) for all infants whose age at death was known (n=144). Twenty-nine infants with a single cardiac gene variant died at a median age of 10.1 weeks (IQR=8.9, 13.0), whereas the 14 infants that were found to have a single variant in a gene related to neurologic function died at a median age of 16.1 weeks (IQR=11.0, 21.3). The 11 infants with a single variant in a ROS pathway gene died at a median age of 12.7 weeks (IQR=9.4, 21.4). ANOVA was run in R and revealed that these differences were significant (P=0.053).

## Discussion

We set out to explore the potential contribution of gene variants to the etiology of SUID. We identified numerous variants in genes associated with SUID, remarkably in 63.4% of our cohort. Some of these are variants known for their involvement in cardiac diseases, neurological disorders, the regulation of neuronal excitability and the response to hypoxia and oxidative stress (ROS). Gene enrichment studies in affected infants further confirmed the validity of these findings.

## ROS Pathway Variants

Each of the genes in the ROS pathway in which P/LP variants were identified contributes to the regulation of the response to hypoxia, in particular the activation of HIF1α and the production of ROS (Yuan, 2013). For example, *AAGAB,* highly expressed in the brain, regulates *PTEN* nuclear translocation to promote the functional neuronal recovery following hypoxic-ischemic induced brain injury (Dai, 2021). *ATM* is activated by hypoxia and re-oxygenation and thus acts as a redox sensor (Hammond, 2004). It plays a critical role in maintaining cellular redox homeostasis (Hammond, 2004). Dysfunction in *ATM* has been implicated in heart failure and sudden death. *BRCA1* plays a major role in the hypoxic response by regulating HIF-1α stability, and also rescues neurons from cerebral ischemia/perfusion injury through the NRF2 signaling pathway (Xu, 2018). *CFTR* is well studied in cystic fibrosis – and associated with respiratory failure, but *CFTR* also functions as a regulator of ROS and has been associated with impairment in oxidative phosphorylation (Ntimbane, 2009). *CFTR* belongs to the ATP binding cassette (ABC) transporter superfamily and is also widely expressed in the CNS (Vasiliou, 2009). *COL7A1* is also a regulator in the hypoxic response and HIF-1α, and it is activated by NF-kB (Amelio, 2018). *ITGB3* is required for sustained TGF-β pathway activation (Sesé, 2017). It is activated by HIF1α in hypoxia and its knockdown has been associated with increased apoptosis and reduced neuronal survival and migration, particularly under hypoxic conditions (Sesé, 2017). *ITGB3* is also associated with acute coronary syndromes (Damar, 2020). HIF1α activates the *TGFβ*-*SMAD3* pathway, and both play a critical role in the hypoxic response and response to injury (Basu, 2011). *SMAD3* is also regulated by serotonin (Chen, 2014) which has been implicated in SUID (Cummings, 2019). Variants in this gene have been described in patients that died of sudden cardiac death, heart failure and aortic aneurysm (Loeys, 2018; Engström, 2020; Ou, 2020; Hanna, 2021; Humeres, 2022).

## Potential genetic contributors to SUID vulnerability

Our findings provide further evidence that genetic markers are important indicators for SUID and may contribute to their vulnerability as hypothesized in the triple risk hypothesis (Guntheroth, 2002). The number of genetic variations associated with cardiac and neurological disturbances was striking and could contribute to an increased vulnerability during the autoresuscitation response, which critically depends on functional cardiorespiratory coupling and the generation of a hypoxic response. An important consideration that emerges from this and prior genetic studies are the implications for later stages of life. Most individuals with these variants will likely survive the critical period that characterizes SUID but may be at risk for sudden death later in life when faced with other stressors or conditions. For example, we found variants in *SCN1A* and *SCN8A*, genes which are known to be associated with SUDEP and have been previously described in SUID, though recognizable seizures had not been observed in these individuals (Kalume, 2013; Hammer, 2016; Chahal, 2020; Rochtus, 2020; Chahal, 2021; Halvorsen, 2021; Keywan, 2021; Koh, 2022). We also found many variants in genes associated with cardiac disorders linked to sudden death. For example, Long QT syndrome (*CACNA1C*, *KCNH2*) which has been previously implicated in SUID, though these disorders had not yet been observed in these infants (Sutphin, 2016; Baruteau, 2017; Tester, 2018; Chahal, 2020; Moore, 2020; Farrugia, 2021; Halvorsen, 2021; Keywan, 2021; Koh, 2022). In addition, we identified affected infants with variants previously reported in ClinVar to be LP/P for conditions such as blood disorders (*ALB, HBA2*) that have been associated with fatal outcomes, but which were not diagnosed in the infants in which these variants were found (Toye, 2012; Russo, 2019). These blood disorder variants are especially notable because transition from fetal hemoglobin to that of adults has been implicated in SIDS (Blix, 2019). Adult hemoglobin, which has a lower oxygen affinity, replaces fetal hemoglobin postnatally over a period of six months (Oski, 1979). Variants associated with deafness (*GPR98/ADGRV1*) are also notable, since inner ear defects have previously been implicated in some infants that succumbed to SUID (Rubens, 2008; Ramirez, 2016). Several of the variants found in our study were previously reported to be pathogenic for disorders of the muscles including spasticity or parapalegia, (*C12orf65/MTRFR*, *DYNC1H1, PIEZO2, SPG7*). Gene enrichment studies aligned with these findings in that notable enrichments in affected infants included motor activity (GO:0003774). Some of these disorders have been associated with respiration dysfunction which could contribute to an individual’s vulnerability.

We found many genes associated with the regulation of the hypoxic response and ROS production as mentioned above. HIF-1α and ROS are produced under conditions of chronic intermittent hypoxia (Garcia, 2016), conditions that are seen in apnea of prematurity (Gauda, 2018) and in recurrent apneas thought to precede SUID in some infants. ROS production is a major driver of dysautonomia and could contribute to an abnormal hypoxic response and failed arousal. Disturbances in the regulation of HIF1α and ROS production could exaggerate the accumulation of ROS in response to conditions like intermittent hypoxia thereby increasing the vulnerability of the child to succumb to an exogenous stressor experienced e.g. under prone conditions. This is consistent with pathological discoveries made in children that succumbed to SIDS (Rognum, 1991; Poulsen, 1993; Stoltenberg, 1993). As previously discussed, the identification of a variant is not sufficient to predict how a variant affects the hypoxic response at the neuronal level, let alone at the level of the child (Opdal, 2004). Follow-up studies that generate these human variants in animal models or in IPSCs or human organoids could provide important additional mechanistic clues. Without these functional insights, these discoveries provide novel evidence that aside of the known cardiac, neurological, and metabolic genes, it will be important to consider also genes associated with the regulation of the hypoxic response as potential biomarkers for SUID. However, at this point it is too early to consider certain variants or groups of variants as biomarkers for SUID.

Statistical analysis of genes and pathways provided novel biological insight. For example, the *F2R* gene contained a splice site variant that was detected in six infants, including both individuals of European and African descent, but only one healthy adult. While primarily a coagulation factor, *F2R* has also been found to block neuronal apoptosis (Guo, 2004), which has been implicated in SUID, particularly in the medulla (Ambrose, 2019). Variants in both *F2R* and *HSF1* that were more frequent in affected infants than healthy adults were responsible for the significance of GO:0043280 (positive regulation of cysteine-type endopeptidase activity involved in apoptotic process) in our analysis. Another finding of note was that SUID infants were more likely than the healthy adults to have functional variants in mucin genes. Disruptions to mucin genes could lead to pathogen susceptibility and/or respiratory difficulties (Pinzón, 2019). These variants found through statistical analysis, however, were not included here because they did not pass our rigorous filtering, which excluded all variants found in healthy adults. The absolute exclusion of these variants might not necessarily make sense when investigating such a complex disorder as SUID. The triple risk hypothesis suggests that there will be many infants with similar genetic vulnerabilities, not exposed to an exogenous stressor, that will survive infancy, so perhaps excluding variants found in healthy adults or that are found more commonly in a population is not the best approach when examining these infants. Another example of this is that in our cohort, 36 children (24.8%) were found with variants in genes involved in serotonin binding and reception which have been implicated previously in SUID (Haynes, 2023). However, none of these variants made it past our filtering mostly because they were found more frequently in the overall population and thus excluded, but it is possible that some of these variants could have contributed to the overall vulnerability of the infant. This filtering approach is a possible limitation of this study.

## Study Limitations

We were able to establish demographic aspects such as race/ethnicity, biological sex, and age at death for all but a small proportion of the cohort. For the majority of infants, 87.9%, there were at least partial ME notes describing details surrounding the death scene and/or autopsy results. These details were used to determine the extrinsic and vulnerability risk factors for each infant. However, we were not able to completely characterize the cohort regarding attributes such as the presence of recent illness, medical history, environmental exposures, or even the sleeping circumstances at the time of death due to the partial nature of the notes. The autopsies for 103 of the infants (71.0% of the cohort) received from the NIH NBB were performed by the same medical examiner. For 19 of these, there were no notes beyond race, sex, age at death, and cause of death. This information helps to identify important phenotypic characteristics of an individual and gives investigators insight into their genetic profile. Detailed phenotypic characteristics are highly valuable for establishing a potential causation regarding the genotype of an individual. In addition, parental data was not available for this analysis, and variants were assumed to be *de novo*. Analysis of trios where this could be confirmed would further strengthen the study.

Mismatched racial proportions between the healthy adults and affected infants are another possible limitation. For the gene level analysis, we demonstrated that significant genes were not confounded by racial differences in the number of variants present (Figure 5). At the variant level, this discrepancy was addressed by matching the appropriate allele frequency in gnomAD specific to the race/ethnicity of the infant in which candidate alleles were identified.

## Conclusion

The present study is one of the first whole genome sequencing studies of infants that died suddenly and unexpectedly. Consistent with the triple risk hypothesis, we find numerous gene variants that could contribute to an increased vulnerability to sudden death. Interestingly, many of those genes have previously been implicated in other causes of sudden death including cardiac death or sudden death in epilepsy. Thus, we hypothesize that many children will survive the critical time period associated with SUID. They will carry this vulnerability into adult life, when they might face other stressors for example those known to be associated with sudden cardiac death. This highlights the importance of recognizing vulnerability early in life, which may not only help to prevent SUID, but sudden death in general.

## Acknowledgments

We would like to thank John and Heather Kahan for their great efforts, financial support, and inspiration to conduct this study.

## Author Contributions

Angela M. Bard: conceptualization, data curation, formal analysis, investigation, methodology, project administration, supervision, validation, visualization, writing-original draft, writing-review & editing. Lindsay V. Clark: conceptualization, data curation, formal analysis, investigation, methodology, software, supervision, validation, visualization, writing-original draft, writing-review & editing. Erdal Cosgun: conceptualization, data curation, formal analysis, investigation, methodology, software, supervision, validation, writing-review & editing. Kimberly A. Aldinger: conceptualization, formal analysis, investigation, methodology, supervision, validation, writing-review & editing. Andrew Timms: data curation, formal analysis, investigation, software, supervision, validation, writing-review & editing. Lely A. Quina: investigation, methodology, validation. Juan M. Lavista Ferres: investigation, resources, software, validation. David Jardine: investigation, methodology, validation. Elisabeth Haas: investigation, methodology, validation. Tatiana M Becker: project administration, validation. Chelsea M. Pagan: project administration, validation. Avni Santani: data curation, validation, writing-review & editing.

Soumitra Barua: data curation, validation. Diego Martinez: data curation, validation. Zakkary McNutt: data curation, validation. Addie Nesbitt: data curation, validation. Edwin Mitchell: validation, writing-review & editing. Jan-Marino Ramirez: conceptualization, formal analysis, funding acquisition, investigation, resources, supervision, validation, visualization, writing-original draft, writing-review & editing.

## Data Availability

The data that support the findings of this study are available on request from the corresponding author. The data are not publicly available due to privacy or ethical restrictions.

